# Antidepressant switching as a proxy phenotype for drug non-response: investigating clinical, demographic and genetic characteristics

**DOI:** 10.1101/2024.11.09.24316987

**Authors:** Chris Wai Hang Lo, Alexandra C. Gillett, Matthew H. Iveson, Michelle Kamp, Chiara Fabbri, Win Lee Edwin Wong, Dale Handley, Oliver Pain, Evangelos Vassos, Naomi R. Wray, Heather C. Whalley, Danyang Li, Allan H. Young, Andrew M. Mcintosh, Cathryn M. Lewis

**Author notes:** **Correspondence** Professor Cathryn M. Lewis, Social, Genetic and Developmental Psychiatry Centre, Institute of Psychiatry, Psychology and Neuroscience, King’s College London, Memory Lane, London, United Kingdom, SE5 8AF.

## Abstract

**Background:** Selective serotonin reuptake inhibitors (SSRIs) are a first-line pharmacological therapy in major depressive disorder (MDD), but treatment response rates are low. Clinical trials lack the power to study the genetic contribution to SSRI response. Real-world evidence from electronic health records provides larger sample sizes, but novel response definitions are needed to accurately define SSRI non-responders.

**Methods:** In UK Biobank (UKB) and Generation Scotland, SSRI switching was defined using a ≤ 90-day gap between prescriptions for an SSRI and another antidepressant in primary care. Non-switchers were participants with ≥ 3 consecutive prescriptions for an SSRI. In UKB, clinical, demographic and polygenic score (PGS) associations with switching were determined, and the common-variant heritability was estimated.

**Results:** In UKB, 5,133 (13.2%) SSRI switchers and 33,680 non-switchers were defined. The mean time to switch was 28 days (IQR: 17-49). Switching patterns were consistent across UKB and Generation Scotland (n = 498 switchers). Higher annual income and educational levels (OR [95% CI] for university degree: 0.73 [0.67-0.79], compared to no qualifications) were associated with lower levels of switching. PGS for non-remission, based on clinical studies, were associated with increased risk of switching (OR: 1.07 [1.02-1.12], p=0.007). MDD PGS and family history of depression were not significantly associated with switching. Using GCTB, the heritability of SSRI switching was approximately 4% (SE: 0.016) on the observed scale.

**Conclusion:** This study identified SSRI switching as a proxy of non-response, scalable across biobanks with EHR, capturing demographic and genetics of treatment non-response, and independent of MDD genetics.

## Introduction

Worldwide, approximately 300 million people suffer an episode of major depressive disorder (MDD) during their lifetime (1). Since the 1990s, selective serotonin reuptake inhibitors (SSRIs) have shown comparable efficacy for MDD treatment in primary care settings, and have become one of the pharmacological treatment options along with tricyclic antidepressants (TCAs) (2). SSRIs have gained in popularity and become the first-line pharmacological treatment in MDD based on safety profiles (3,4). However, considerable variability exists in antidepressant response (5), with only about one-third of antidepressant users achieving clinical remission with their first prescribed antidepressant (6). Another third of patients go on to develop treatment-resistant depression (TRD), defined as the lack of response to two antidepressants with adequate duration and dosage (7). Identification of factors that predict response and non-response to antidepressants would enable personalised prescribing and improve treatment for MDD.

Multiple factors have been associated with response and non-response to antidepressant treatment. For example, childhood trauma is associated with poorer response to antidepressants (8) and, in clinical trials, higher body mass index (BMI) and neuroticism scores were significantly associated with antidepressant response (9,10). Patients with TRD often have higher depression symptom severity (11,12), and observational studies suggest that TRD is correlated with sociodemographic characteristics, such as unemployment (12). However, baseline MDD severity was not associated with symptom-level response in a meta-analysis of 91 clinical trials (13). Biomarkers for antidepressant response, including brain-derived neurotrophic factor (14), cortisol (8) and inflammatory markers (15), show inconsistent results.

Genetic factors have been associated with antidepressant response. Cytochrome P450 variants play a minor role in adverse events and response by affecting metabolism of antidepressants (16–19). The largest genome-wide association study (GWAS) of clinical studies to date showed antidepressant remission was heritable, with a single-nucleotide polymorphism (SNP)-based heritability of upto 40% (20). However, genetic studies performed with clinical trials are under-powered to discover SNPs at genome-wide significance. Stringent inclusion criteria may also limit the generalisability of genetic findings to a population-wide level. Other study designs are therefore required to increase the power to identify the genetic component of antidepressant response. Real-world data from electronic health records (EHR) could fill this gap, since large sample sizes are available in biobanks with genetic data.

In EHR, defining treatment response phenotypes is challenging, as response (or resistance) to antidepressant treatment is not directly coded in most records. Using clinical records, proxy phenotypes can be captured from unstructured text using natural language processing algorithms (21). Alternatively, phenotypes can be defined from structured prescription records, which are more readily available and scalable in population-wide biobanks. One feasible strategy is to capture switching events between antidepressants as an indication of non-response (22). This approach reflects clinical guidelines, where patients who fail to respond to an antidepressant are recommended to switch to a different drug. Antidepressant switching in EHR has been used to define TRD, where two switches occur within a single episode of depression (23). Antidepressant switching is also used in clinical trials as an alternative therapeutic strategy following inadequate response to the first antidepressant (often an SSRI) (24,25).

In this study, we use primary care prescribing records in UK Biobank and dispensing records in Generation Scotland (26), to define a phenotype of switching from an SSRI to another antidepressant (of any class) within an episode of depression. We characterize the prescription patterns, and investigate the clinical, demographic, and polygenic predictors of SSRI switching. We further perform GWAS of SSRI switching, showing that switching is heritable, and we propose switching as a proxy measure of non-response to SSRIs.

## Methods and Materials

### Primary Sample – UK Biobank (UKB)

UKB is a prospective health study that recruited over 500,000 volunteers aged 40 to 69 years in the United Kingdom from 2007 to 2010 (27,28). Genome-wide genotyping, available for all UKB participants, underwent standard quality control (QC) and imputation. Further description of UKB samples, as well as details on genotyping QC and imputation, is available in **Supplementary Methods,** and an analysis flowchart is shown in **Figure 1**. Linkage with primary care data is available for ∼230,000 UKB participants, containing clinical events (coded by READ v2 or CTV-3) and prescription records (coded by READ v2, READ v3, BNF or dm+d) (29). READ v3 diagnosis codes used are listed in **Supplementary table 1**, with antidepressants and mapped drug classes listed in **Supplementary table 2**.

**Figure 1.**
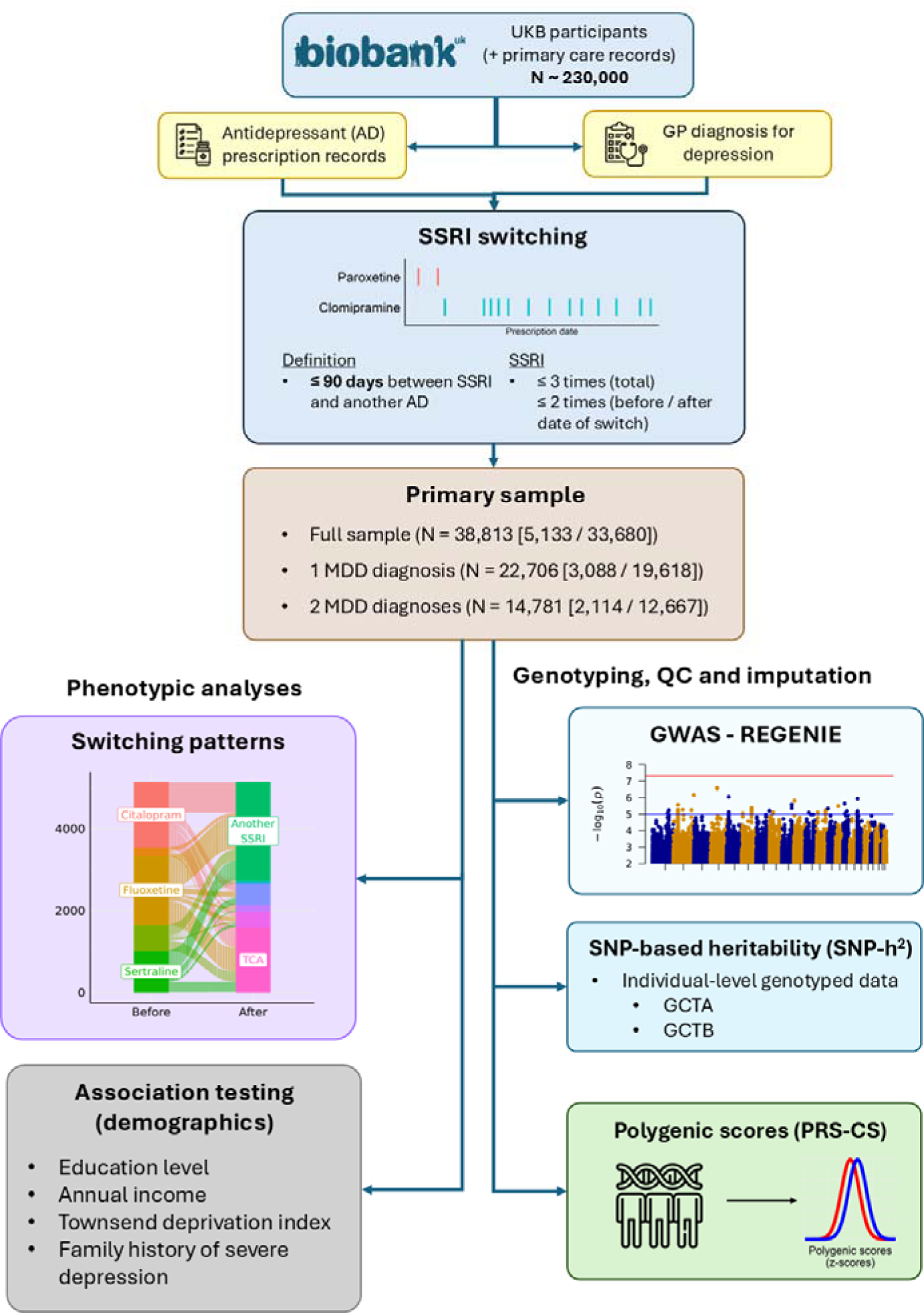
Study design overview for primary sample (UK Biobank) **Legends** Icons used created by Team Iconify, DailyPm Studio and Freepik from Flaticon. **Abbreviations** GCTA = Genome-wide Complex Trait Analysis; GCTB = Genome-wide Complex Trait Bayesian; GWAS = genome-wide association study; QC = quality control; SNP = single nucleotide polymorphism; SSRI = selective serotonin reuptake inhibitors; UKB = UK Biobank.

Patients with depression were identified using primary care diagnosis records for depressive disorders, using a previously validated algorithm (23). Participants with any primary care diagnosis for bipolar disorder, psychosis or substance abuse were excluded. Not all patients prescribed SSRIs had a depression diagnostic code assigned. To examine the impact of diagnosis, we analysed three datasets of participants prescribed an SSRI: (1) all patients satisfying the switcher / non-switcher criteria regardless of depression diagnoses; (2) patients with ≥ 1 depression diagnostic record; and (3) participants with ≥ 2 depression diagnostic records.

Sociodemographic and clinical variables extracted from UKB were self-reported sex (field ID 31-0.0), educational qualifications (field ID 6138-0.0, with “none of the above” as reference level), annual income (as average total household income before tax, field ID 738-0.0, with “less than £18,000” as reference level), Townsend Deprivation Index (field ID 189-0.0), BMI (field 21001-0.0) and family history of depression (with “no” as reference level). A positive family history of depression was defined where at least one parent (field ID 20107-0.0 and 20110-0.0) or sibling (field ID 20111-0.0) reported severe depression. Polygenic scores (PGSs) for MDD (UKB participants removed) (30), schizophrenia (31) and antidepressant non-remission (20) were computed using PRS-CS (32) with the GenoPred pipeline (version 1) (33,34). Details of polygenic scoring methods and GWAS summary statistics are summarised in **Supplementary Methods** and **Supplementary table 3.**

### Replication sample – Generation Scotland

Replication analyses were performed in Generation Scotland to compare switching patterns across healthcare practices. Generation Scotland is a family-based longitudinal study which recruited over 24,000 volunteers from 2006 to 2011, with information on demographics, physical and mental health measurements (26). Linkage to dispensed prescriptions in primary care was available for over 90% of participants (26). Details on Generation Scotland replication sample are summarised in **Supplementary Methods**.

### Phenotype definition – SSRI switching

SSRI switchers were defined as participants being prescribed an SSRI who then received a prescription for another antidepressant within a 90-day window, from 5-95 days of the initial prescription (a *switching* event). The following additional criteria were applied:

1. a minimum of 5-day window between prescriptions to avoid capturing overlapping prescriptions of two antidepressants (augmentation) as switches;
2. the pre-switch SSRI was prescribed ≤ 3 times in all prescribing history to ensure it was not prescribed in future treatment episodes;
3. the pre-switch SSRI was prescribed ≤ 2 times before the switch date, to capture early switchers specifically; and
4. the pre-switch SSRI was prescribed ≤ 2 times after the switch date, to ensure augmentation was not captured, while giving a brief allowance period for cross-tapering.

SSRI non-switchers were defined as patients who did not switch from any SSRIs, and received ≥ 3 prescriptions for an SSRI.

The SSRI index date was defined as the first prescription date for the SSRI in switchers and non-switchers. A schematic figure of definitions of switchers and non-switchers is shown in **Supplementary figure 1**.

### Analysis of switching patterns, clinical and demographic variables, polygenic scores

Descriptive analyses were performed on SSRI switchers and non-switchers, by index SSRI drug, drug class post-switch, time to switch and age at index date. Differences in the distribution of the variables between SSRI switchers and non-switchers were assessed by nonparametric statistical tests, including Pearson’s chi-square test (for binary variables), Kruskal-Wallis rank-sum test (for categorical variables of more than two levels), and Wilcoxon rank-sum test (for continuous variables).

Associations between switching and sociodemographic variables at baseline assessment and PGSs were tested by logistic regression, with models adjusted for self-reported sex (with female as reference level), index date of SSRI, and assessment centre (with “Centre_10003” as reference level). Associations with PGSs were further adjusted for 10 principal components for population stratification. For related individuals, one participant was removed based on third-degree relatedness (kinship coefficient < 0.044) by greedy matching, with cases being preferentially retained. Statistical significance was assessed by likelihood ratio test and corrected for multiple testing by Bonferroni correction within each sample (p ≤ 0.0071, correcting for 4 sociodemographic variables [educational qualifications, annual income, family history of severe depression and Townsend Deprivation Index] and 3 PGSs [non-remission, MDD, schizophrenia]).

### Genome-wide association study

Genome-wide association analysis was performed on SSRI switching using REGENIE (35), a two-step software for genome-wide analysis. Given the low ratio of switchers / non-switchers, SNPs with low allele frequencies might go into quasi-complete separation when applying standard logistic regression models in GWAS (35). Therefore, SNP effect sizes underwent Firth correction to control for false positives as recommended in REGENIE documentation (35).

For genetic analyses, we tested for differences in the distributions of assessment centre and genotyping batch between switchers and non-switchers by Kruskal-Wallis rank sum test, to assess which covariates to be included (see **Supplementary table 9**). Covariates included in the GWASs were SSRI index date, genetic sex, assessment centre and ten principal components for population stratification.

### SNP-based heritability (*h^2^*) estimation

SNP-based heritability was estimated using two genomic relatedness–based restricted maximum likelihood (GREML)-based methods, Genome-wide Complex Trait Analysis (GCTA) (36) and Genome-wide Complex Trait Bayesian (GCTB) (37). SNP-based heritability (*h^2^*) was reported on the observed scale since the sample was unselected for SSRI treatment. In GCTB, the *h^2^* estimates were constrained to be between 0 and 1 in each iteration, and the distributions of *h^2^* estimates can be skewed when the true *h^2^* is close to 0. To strengthen the robustness of our findings, the posterior mode and 95% highest posterior density (HPD) credible intervals were also reported for GCTB. The degree of polygenicity (*Pi*) and negative selection (*S*) were also reported. Full details of *h^2^*estimation are available in **Supplementary methods**.

## Results

### Descriptive analyses

In UKB, a total of 5,133 SSRI switchers and 33,680 non-switchers were identified from prescription records (full sample). Baseline characteristics of SSRI switchers and non-switchers are given in **Table 1 and Supplementary figure 2**. Patients in the full sample had a median of 18 SSRI prescriptions (IQR: 7-47; **Figure 2, Supplementary table 4**) across all prescribing history, primarily spanning from the 1990s to 2018 (**Supplementary figure 3**). Of these participants, 3,088 (60%) switchers and 19,618 (58%) non-switchers had at least one diagnostic record for MDD, with 2,114 (41%) switchers and 12,667 (38%) non-switchers having at least two MDD diagnostic records (**Supplementary figure 2)**. In the full sample, 67% were female, and 96% were of white ethnicity (**Table 1**). Approximately half of participants had at least one prescription for a tricyclic antidepressant (TCA) (full sample: *N* = 18,125 [46.7%]; one depression record: *N* = 11,320 [49.9%]; two depression records: *N* = 7,749 [52.4%]; **Supplementary table 5**), and 13-18% received ≥ 1 prescription for a serotonin-norepinephrine reuptake inhibitor (SNRI) (**Supplementary table 5**). In Generation Scotland, a total of 498 SSRI switchers and 1,279 non-switchers with at least one diagnostic record for depression were identified (**Supplementary table 7**).

**Figure 2.**
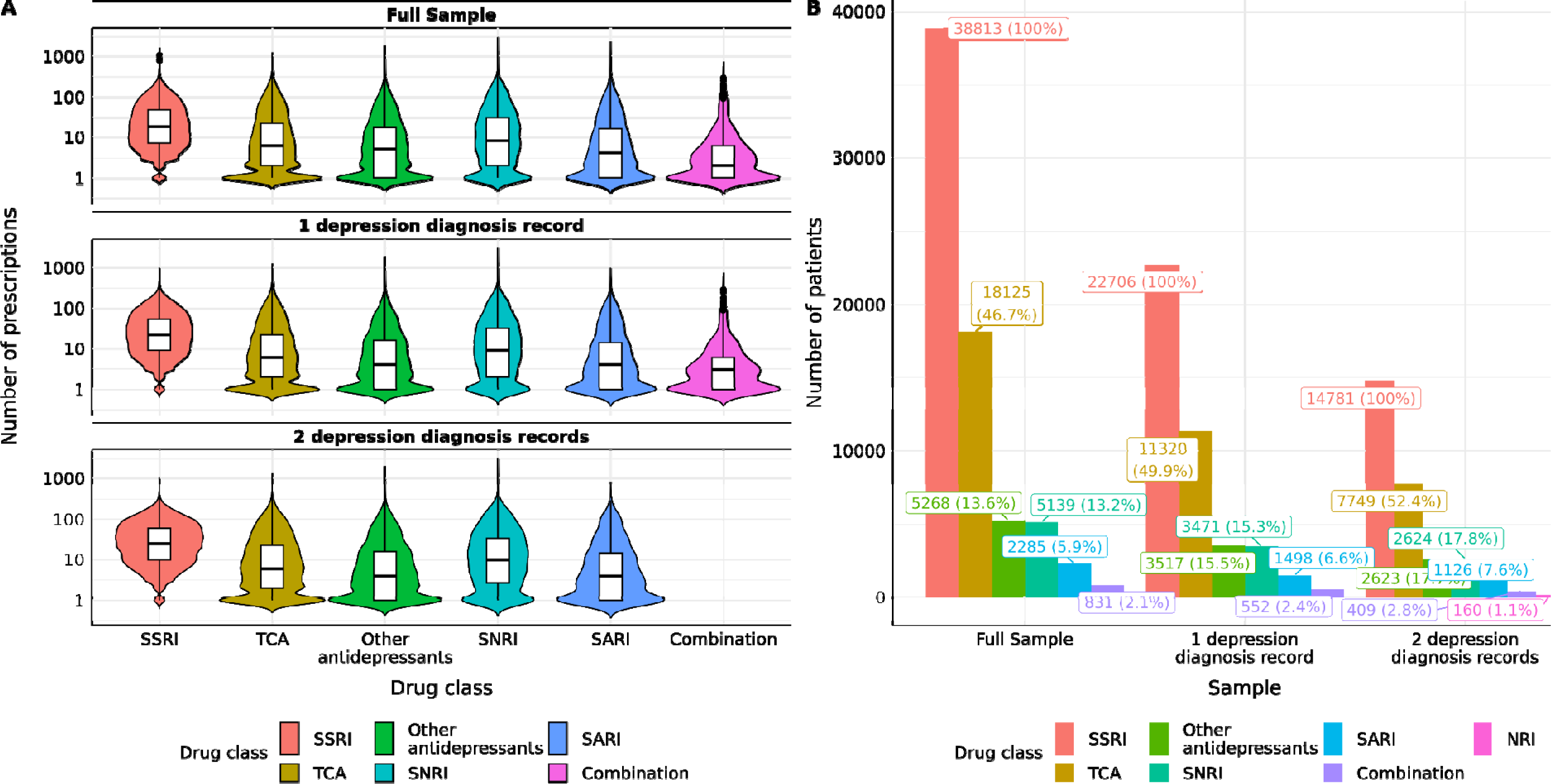
Number of antidepressant prescriptions in primary sample (UK Biobank) **Caption** (A) Number (Median [IQR]) of prescriptions of primary sample (by drug classes with number of patients > 500); (B) Proportion of patients receiving at least 1 prescription for a particular drug class. Only drug classes consisting of > 1% of sample sizes were labelled. Details on the statistics for both figures were available in supplementary materials. **Abbreviations** MAOI = monoamine oxidase inhibitors; NDRI = norepinephrine-dopamine reuptake inhibitors; NRI = norepinephrine reuptake inhibitors; SARI = serotonin antagonist and reuptake inhibitors; SNRI = serotonin–norepinephrine reuptake inhibitors; SSRI = selective serotonin reuptake inhibitors; TCA = tricyclic antidepressants.

**Table 1.**
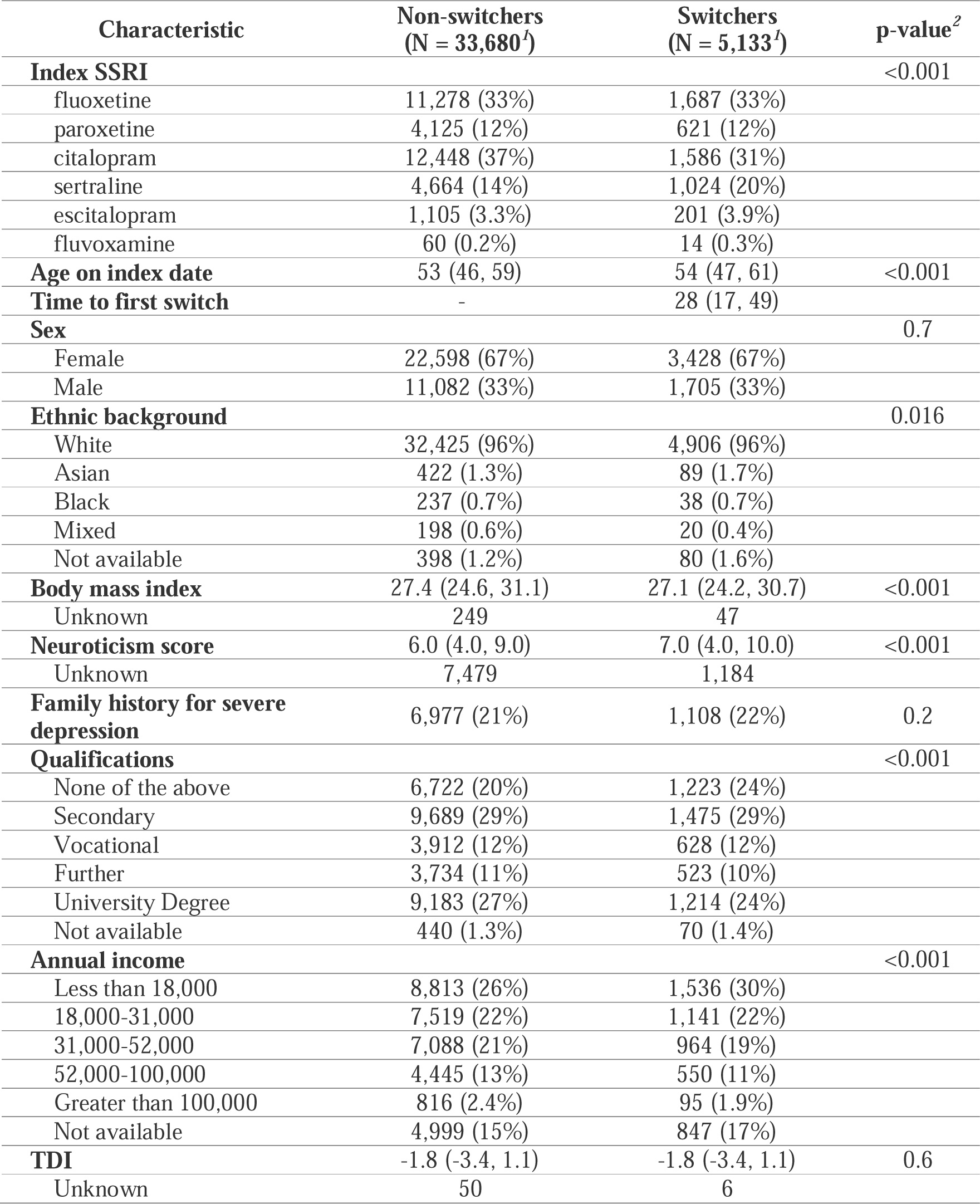
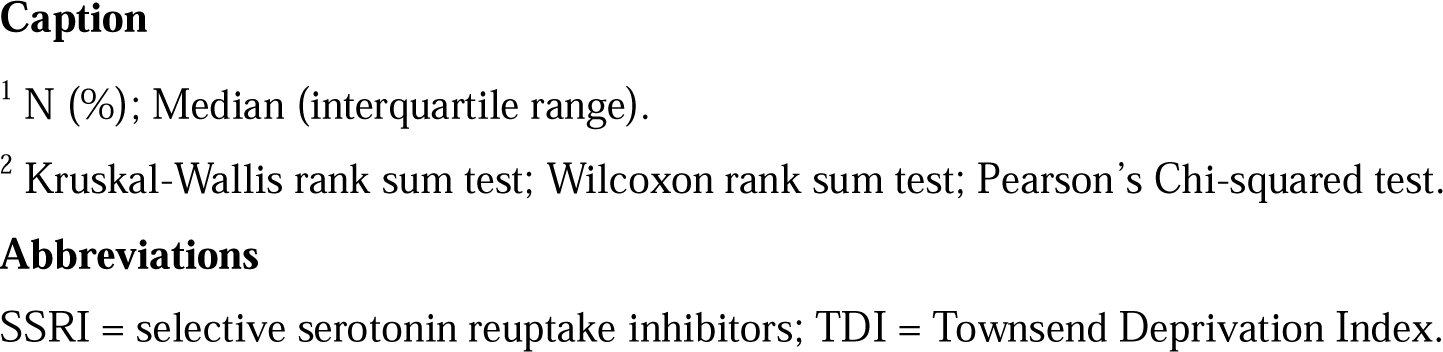
Primary sample (UK Biobank) summary.

### Patterns of SSRI switching

In both UKB and Generation Scotland, the most prescribed/dispensed SSRI antidepressants were fluoxetine and citalopram. Around one-third received fluoxetine (UKB [**Table 1**]: N = 1,687 [33%] switchers and 11,278 [33%] non-switchers; Generation Scotland [**Supplementary table 7**]: N = 165 [33%] switchers and 433 [34%] non-switchers). Another one-third received citalopram (UKB: N = 1,586 [31%] switchers and 12,448 [37%] non-switchers; Generation Scotland: N = 190 [38%] switchers and 604 [47%] non-switchers). Paroxetine and sertraline were also commonly prescribed/dispensed, to over 10% of participants each.

Most SSRI switches occurred within six weeks of the index prescription in both UKB (median time to switch in days [IQR]: 28 [17-49], **Table 1**) and Generation Scotland (31 [31-61], **Supplementary table 7**). Distributions of time to switch were similar across index SSRIs in UKB (**Supplementary figure 4**). The proportion of switchers and time to switch were generally comparable across sex (**Supplementary figure 6**). In UKB, approximately half of switching events were to another SSRI (N = 2,380; 46%), or a TCA (N = 1,597; 31%) (**Supplementary table 8, Figure 3**). Similar patterns were observed in Generation Scotland, with half switching to another SSRI (N = 237; 47.6%), with fewer patients changing to TCA after switching (N = 73; 14.7%) (**Supplementary figure 7, Supplementary table 9**).

**Figure 3.**
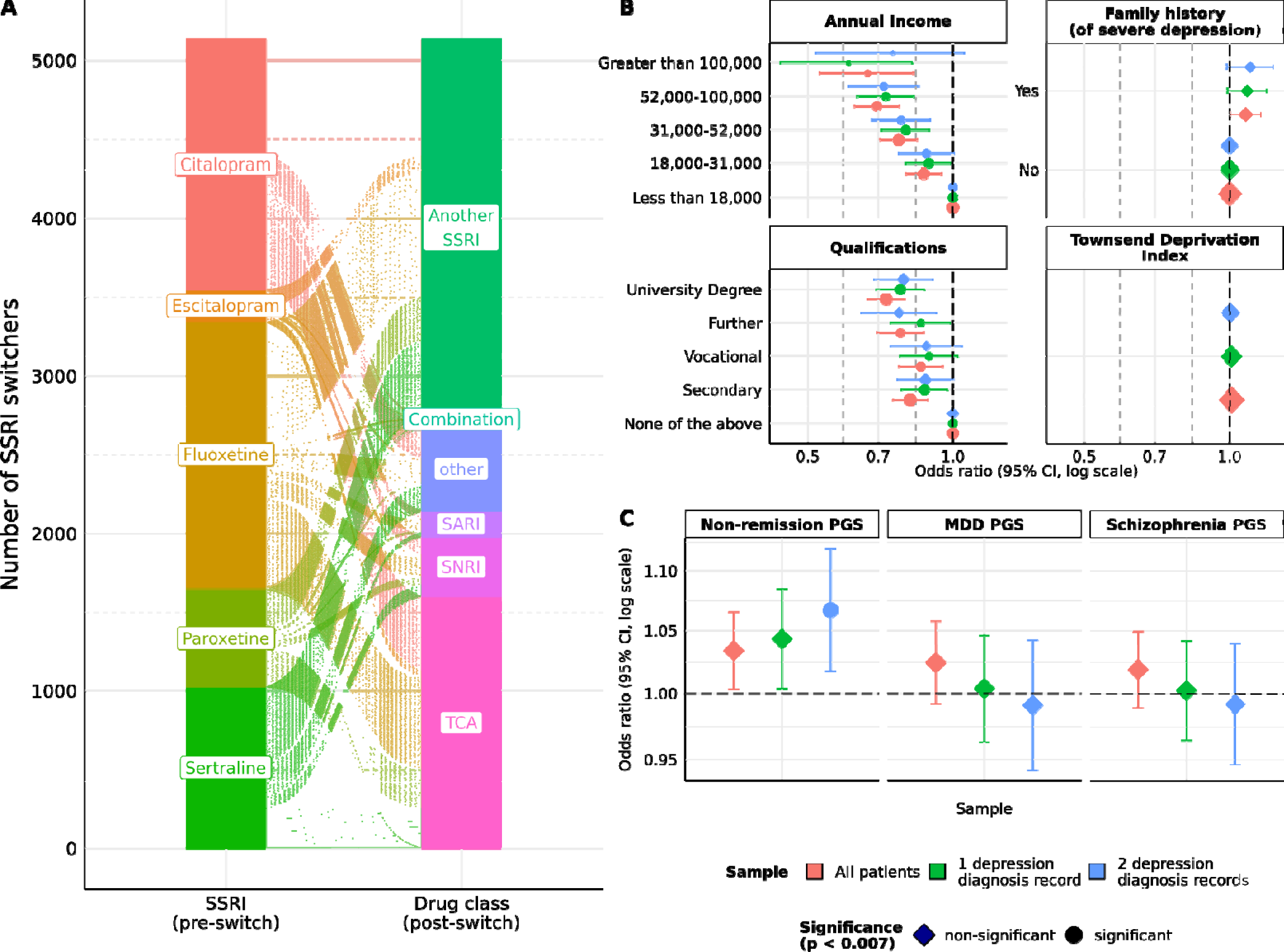
Switching patterns and association testing with clinical, sociodemographic variables and polygenic scores in primary sample (UK Biobank) **Caption** (A) SSRI switching patterns in UK Biobank, stratified by index SSRI and drug class after switch; (B) Association testing between demographic variables and SSRI switcher statuses; (C) Association testing between polygenic scores and SSRI switcher status. Only SSRIs (pre-switch) and drug classes (post-switch) with at least a sample size of 70 are labelled. **Abbreviations** MDD = major depressive disorder; PGS = polygenic scores; SARI = serotonin antagonist and reuptake inhibitors; SNRI = serotonin– norepinephrine reuptake inhibitors; SSRI = selective serotonin reuptake inhibitors; TCA = tricyclic antidepressants.

Across the 22 assessment centres in UKB, the proportion of switchers differed significantly, varying between 9-19% (p = 2.5e-08; **Supplementary table 10, Supplementary figure 8).** The rate of switching increased with more recent index dates (Pearson correlation coefficient between switching rate and time [*r*] = 0.43, p = 1.0e-05), and the period between index date and switching was shorter (*r* = -0.42, p = 1.5e-05) (**Supplementary figure 9**).

### Associations with sociodemographic variables and polygenic scores

In UKB, higher educational levels were associated with lower odds of SSRI switching (odds ratio for university degree [OR, 95% CI]: 0.73 [0.67-0.79], p = 1.53e-10, compared to the reference group of no qualifications) (**Supplementary table 11**). The results were consistent when samples were limited to at least two MDD diagnoses in primary care (0.79 [0.69-0.91], p = 0.013) and at least one MDD diagnosis (0.78 [0.69-0.87], p = 0.001). Similar findings were observed in annual income, with higher income associated with lower risks of SSRI switching for annual income > £100,000 compared to the reference group of <£18,000 in the full sample (0.66 [0.53-0.83], p = 6.79e-15). Effect sizes were similar when constrained to at least two MDD diagnosis records (0.75 [0.52-1.06], p = 1.43e-04) and one MDD diagnosis record (0.61 [0.44-0.82], p = 4.92e-07). SSRI switching was not associated with Townsend Deprivation Index, a measurement of material deprivation.

Family history of severe depression was only nominally associated with SSRI switching status in the full sample (OR [95% CI]: 1.08 [1.00-1.16], p = 0.048), and was not associated with switching in patients with one (1.09 [0.99-1.19], p = 0.084) or two (1.10 [0.99-1.23], p = 0.088) MDD diagnosis records. The PGS for MDD was not associated with SSRI switching (1.02 [0.99-1.06], p = 0.138, for full sample). Similar results were found when restricting samples to participants with at least one (1.00 [0.96-1.05], p = 0.848) and two (0.99 [0.94-1.04], p = 0.73) MDD diagnostic records.

Higher PGS for antidepressant non-remission was associated with an increased risk of SSRI switching, with modest effect sizes. Only the associations in patients with two MDD diagnostic records survived multiple testing corrections (OR [95% CI]: 1.07 [1.02-1.12], p = 0.007). The associations were nominally significant in the full sample (1.03 [1.00-1.07], p = 0.029), and in participants with a single MDD diagnostic record (1.04 [1.00-1.08], p = 0.031), with similar directions of association (**Figure 3, Supplementary table 11).**

### Genetic analyses

A GWAS was performed using REGENIE, on 4,773 SSRI switchers and 31,561 non-switchers on the full sample. The sample sizes were reduced to 2,868 SSRI switchers and 18,360 non-switchers for one MDD diagnosis record, and to 1,967 switchers and 11,853 non-switchers for two MDD diagnosis records. No variants were identified in either analysis at genome-wide significance (p < 5e-8), with Manhattan plots shown in **Supplementary figures 15-17)**. At a suggestive significance threshold (p<1e-5), 21, 25 and 30 independent SNPs were identified with the full sample, as well as samples with one or two MDD diagnoses respectively.

SSRI switching had a SNP-based heritability significantly different from zero on the observed scale in the full sample (GCTA *h^2^*[SE]: 0.0268 [0.016], p = 0.038; GCTB: 0.0242 [0.01], p = 0.005), as well as participants with one MDD diagnosis record (GCTA: 0.0431 [0.0268], p = 0.048; GCTB: 0.0398 [0.016], p = 0.005) (**Figure 4, Supplementary table 12)**. The posterior mode for *h^2^* in GCTB was also different from zero (0.032; 95% HPD credible intervals [0.012-0.065]). *h^2^* estimates were non-significant for patients with at least two MDD diagnoses records in GCTB (0.035 [0.025], p = 0.08) and GCTA (0.0216 [0.0394], p=0.293).

**Figure 4.**
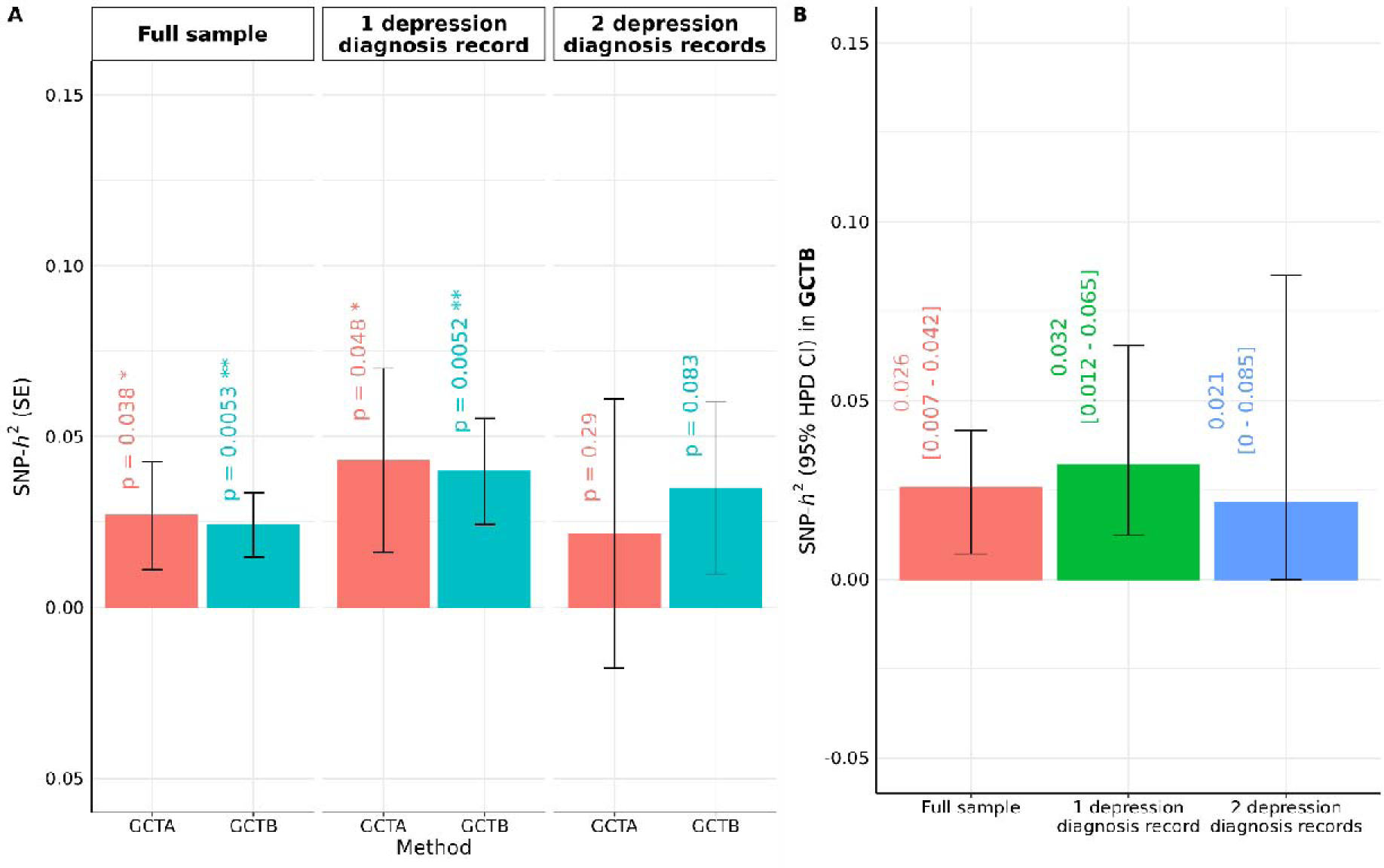
SNP-based heritability estimates for SSRI switching by GCTA and GCTB. **Caption** SNP-based heritability (*SNP-h^2^*) on observed scale stratified by number of depression diagnoses in primary sample, expressed in **(A)** posterior mean (standard error) and **(B)** posterior mode (95% HPD credible intervals) for GCTB. * p-value < 0.05. ** p-value < 0.01. **Abbreviations** GCTA = Genome-wide Complex Trait Analysis; GCTB = Genome-wide Complex Trait Bayesian; HPD = highest posterior probability; SE = standard error; SNP = single nucleotide polymorphism; SSRI = selective serotonin reuptake inhibitors.

## Discussion

EHRs offer promising opportunities to define antidepressant response outcomes from prescribing patterns, but these outcomes have not been well characterised to date. In this study, we used SSRI switching as a proxy phenotype for non-response to antidepressants. The measure reflects the current clinical practice of moving patients from first-line SSRI treatment to a different antidepressant in cases of no- or poor response (3,38), the signs of which could be evident from two to four weeks at the earliest (39). We identify associations of switching with demographic and genetic profiles, and show a modest heritability for the switching phenotype.

Our phenotypic definition of switching in UKB aimed to capture antidepressant switchers, following previous work on over 260,000 participants prescribed antidepressants in the UK Clinical Practice Research Datalink (CPRD) database of primary care records (40). In the CPRD study, most SSRI users switched to another SSRI as second-line therapy (54.1%) (40), which is consistent with our findings. In CPRD, 9.3% of antidepressant users switched, with a median time to switch of 45 days (40). In comparison, SSRI users in UKB had a higher proportion of switching (13.2%), and a shorter time to switch (median: 28 days). The study period for the CPRD study was from 2005 (40), compared to the 1990s in UKB, the period when SSRIs became the first-line therapy in clinical guidelines (2).

We used a 90-day window between prescription dates of two different antidepressants to capture switching events. This contrasts with the shorter window applied in CPRD (40), where switching events were identified from a 30-day or less gap between the expected end dates of the first treatment and the start date of the second treatment. Using longer windows allows us to capture more switchers for genetic analyses, but is less specific to the exact cause of switching in the samples. Of note, using different window lengths did not substantially alter effect sizes for associations between switching and CYP2C19 metaboliser status in a previous UKB analysis (22). Our definition of switching events primarily relies on prescription dates of different antidepressants. This avoids making inferences of treatment duration where it cannot be accurately estimated from dosage instructions and quantity of prescriptions, as in UKB EHR data.

SSRI switching captures demographic and clinical variables associated with non-response to antidepressants. We showed that the proportions of participants switching were lower in those with higher incomes and higher educational levels, which aligns with evidence that higher PGS for education attainment were associated with remission in clinical trials (20). Higher levels of antidepressant response have been associated with higher socioeconomic status (SES) in a systematic review of clinical trials (41), and a Nordic registry study (42). SES was suggested previously to directly contribute to poor prognosis if it was causal to the development of depression itself (43). SES is also correlated with access to treatment and mental health services, which might in turn affect adherence to antidepressant and treatment outcomes (41). Our results confirm that SSRI switching in EHRs shows similar sociodemographic profiles as seen in antidepressant non-response in trials and retrospective clinical studies.

Results from the genetic analyses also support SSRI switching as a proxy phenotype for antidepressant non-response. SSRI switching in UKB was associated with PGS for non-remission, but not with MDD PGS or with a family history of depression. These results indicate that the genetics of SSRI switching overlaps with the genetics of antidepressant response in clinical trials, but is independent of the genetics of MDD (20). MDD PGS captures the genetics of susceptibility and also of symptom severity (44,45), a strong predictor for antidepressant non-response. However, these genetic factors were not correlated with response outcomes in our analysis. Mixed evidence has been found in previous studies, with positive correlations between MDD PGS and poorer response in smaller clinical trials (16,46,47), but none was robust to multiple testing. Our genome-wide analysis of SSRI switching was underpowered to detect specific risk variants, but the identification of this EHR-based phenotype for antidepressant response/non-response opens opportunities for expanding the sample size in other real-world data sources. We sought replication of the UKB SSRI switching phenotype in Generation Scotland, which has 20,000 participants. However, with only 1,777 study members classified as SSRI switchers or non-switchers, limited analysis could be performed. We reported the results here for completeness.

Genetic analyses in UKB revealed a modest heritable signal for SSRI switching (SNP-based *h^2^* = ∼0.04). Antidepressant response was also found to be significantly heritable in a GWAS meta-analysis of clinical trials (20), with estimates of SNP heritability of 0.083 (SE: 0.035) on the observed scale. The lower SNP-based heritability estimates for SSRI switching likely occur because switching is a proxy phenotype which captures non-response alongside adverse events and other features of SSRI treatment. Although genetic correlations cannot be calculated due to limited sample sizes, the positive association between non-remission PGS and SSRI switching confirms a common genetic component for these phenotypes. Our genetic heritability estimates differed between the GREML-based methods of GCTA and GCTB, and stratifying by one or two depression diagnosis records. The *h^2^* estimates with two MDD diagnosis records were not significantly different from zero, meaning we were unable to consistently confirm non-zero heritability across samples, likely due to heterogeneity and insufficient statistical power. However, our results highlight the potential of using switching as a proxy phenotype to capture non-response from antidepressants, which is scalable across EHR data resources (26,48) and will allow future meta-analyses to obtain more robust estimates.

Although SSRI switching appears to capture dimensions of non-response phenotypically and genetically, one should be aware of the heterogeneity in the exact causes of switching in EHR. Patients with MDD could switch antidepressants for a multitude of reasons, including lack of efficacy, non-compliance to treatment and side effects (38). Using dosage information in prescription records would enable further investigation of the causes of switching, yet it is not readily available in UKB prescription records. Stratifying switchers by switching windows might be another feasible strategy, as patients switching earlier are more likely to reflect tolerability issues, while later switchers might be due to a lack of efficacy.

This study had several limitations in data availability and sample sizes in UKB primary care records. Firstly, non-remission PGSs were calculated using genetic studies in antidepressant clinical trials with moderate sample sizes. Using the UKB sample alone, genetic analyses were also underpowered, and at the margins of requirements for robust SNP-based heritability estimates. Larger sample sizes are necessary to replicate the current findings. In GWAS of mental disorders, broadening phenotypic definition increases the power to detect associated loci, but reduces specificity (49,50). We decided to maintain a balance between the two by stratifying SSRI switchers with at least one or two MDD diagnostic records in primary care, to ensure that the antidepressant was prescribed for depression instead of conditions such as anxiety and insomnia (51). However, we did not limit the samples to requiring primary care diagnoses of depression on or before SSRI exposure. As a check, primary care diagnoses of depression preceded index SSRI exposure in over 70% of participants in our samples (**Supplementary table 6**), despite the possible missingness of diagnoses records in EHR. While UKB has rich EHR data on prescribing, it lacks the response measures available in clinical trials, including depression symptom scores at baseline and during treatment. Finally, the choice of treatment and choice to switch treatment, has multiple contributing factors, which our analyses may not have fully accounted for, with the possibility of residual confounding.

## Conclusion

Using primary care records in UKB, we characterised SSRI switching as a data-driven proxy that captures clinical, demographic and genetic dimensions of SSRI non-response. Switching patterns identified in our phenotyping algorithm were consistent with current clinical practice, with most switching to another SSRI. SSRI switching captured the genetics of antidepressant response, and was distinct from the genetics of MDD susceptibility. We also identified a modest, but significant, heritability for SSRI switching. In summary, SSRI switching defined from electronic health records is a valuable phenotype to study the genetics of SSRI response. As a highly scalable phenotype, SSRI switching can contribute to pharmacogenetic research moving towards personalised prescribing.

## Data Availability

TCodes used to create the switching phenotype and perform genetic analyses for this work is publicly available on Github (https://github.com/chrislowh/SSRI_switching). The UK Biobank data was accessed via project 82087. Please visit https://www.ukbiobank.ac.uk/enable-your-research for access. Generation Scotland data are available on reasonable request. Researchers may request access to Generation Scotland data through https://www.ed.ac.uk/generation-scotland/for-researchers.

## Acknowledgements

A preprint of this manuscript is available on Medrxiv at: https://doi.org/10.1101/2024.11.09.24316987. For the purposes of open access, the author has applied a Creative Commons Attribution (CC BY) licence to any Accepted Author Manuscript version arising from this submission. We extend our gratitude to the contributions of other investigators on the “AMBER: Antidepressant Medications: Biology, Exposure & Response” project, including: Edinburgh: Iona Beange, Cristina Douglas, Sue Fletcher-Watson, Mark Somerville, Arlene Casey, Matúš Falis, Mark J Adams, Megan Calnan, Amelia J Edmondson-Stait, David Brici, Anjali K Henders, Quan Nguyen, Sonia Shah, Clara Albiñana and Alicia Walker. We are grateful to all the families who took part in Generation Scotland, the general practitioners and the Scottish School of Primary Care for their help in recruiting them, and the whole Generation Scotland team, which includes interviewers, computer and laboratory technicians, clerical workers, research scientists, volunteers, managers, receptionists, healthcare assistants and nurses.

## Code availability

Codes used to create the switching phenotype and perform genetic analyses for this work is publicly available on Github (https://github.com/chrislowh/SSRI_switching).

## Data sharing statement

The UK Biobank data was accessed via project 82087 — for access, go to https://www.ukbiobank.ac.uk/enable-your-research. Generation Scotland data are available on reasonable request. Researchers may request access to Generation Scotland data through https://www.ed.ac.uk/generation-scotland/for-researchers.

