## Supplementary Materials for "Antidepressant switching as a proxy phenotype for drug non-response: investigating clinical, demographic and genetic characteristics"

Chris Wai Hang Lo^1^, Alexandra C. Gillett^1,2^, Matthew H. Iveson^3^, Michelle Kamp^1^, Chiara Fabbri^4^, Win Lee Edwin Wong^1,5^, Dale Handley^1^, Oliver Pain^6^, Evangelos Vassos^1,2^, Naomi R. Wray^7,8,9^, Heather C. Whalley^3,10^, Danyang Li^1^, Allan H. Young^11,12^, Andrew M. Mcintosh^3,13^, Cathryn M. Lewis^1,2,14^

^1^ Social, Genetic & Developmental Psychiatry Centre, Institute of Psychiatry, Psychology and Neuroscience, King's College London, London, United Kingdom

^2^ National Institute for Health Research Maudsley Biomedical Research Centre at South London and Maudsley NHS Foundation Trust and King’s College London, London, United Kingdom

^3^ Division of Psychiatry, Centre for Clinical Brain Sciences, University of Edinburgh, Edinburgh, United Kingdom

^4^ Department of Biomedical and Neuromotor Sciences, University of Bologna, Bologna, Italy

^5^ Department of Pharmacology, Yong Loo Lin School of Medicine, National University of Singapore, Singapore, Singapore.

^6^ Department of Basic and Clinical Neuroscience, Institute of Psychiatry, Psychology and Neuroscience, King’s College London, London, United Kingdom

^7^ Institute for Molecular Bioscience, University of Queensland, Brisbane, Queensland, Australia

^8^ Queensland Brain Institute, University of Queensland, Brisbane, Queensland, Australia

^9^ Department of Psychiatry, University of Oxford, Oxford, United Kingdom

^10^ Generation Scotland, Centre for Genomic and Experimental Medicine, Institute of Genetics and Cancer, University of Edinburgh, Edinburgh, United Kingdom

^11^ Department of Psychological Medicine, Institute of Psychiatry, Psychology and Neuroscience, King’s College London, London, United Kingdom

^12^ South London and Maudsley NHS Foundation Trust, London, United Kingdom

^13^ Institute for Genomics and Cancer, University of Edinburgh, Edinburgh, United Kingdom

^14^ Department of Medical & Molecular Genetics, King’s College London, London, United Kingdom

**Antidepressant switching as a proxy phenotype for drug non-response: investigating clinical, demographic and genetic characteristics**

[Supplementary table 13. Top 40 hit SNPs with p < 1e-5 for SSRI switching (full sample)	28](#_Toc190278636)

[Supplementary table 14. Top 40 hit SNPs with p < 1e-5 for SSRI switching (≥ 1 MDD diagnosis record)	30](#_Toc190278637)

[Supplementary table 15. Top 40 hit SNPs with p < 1e-5 for SSRI switching (≥ 2 MDD diagnosis records)	32](#_Toc190278638)

### Supplementary Methods

#### Primary Sample – UK Biobank (UKB)

##### Sample description

The UKB is a population-wide prospective cohort study consisting of 488,377 participants recruited from the United Kingdom (UK) (1). All participants provided consent upon assessment, and completed self-assessment questions on sociodemographic and lifestyle characteristics, as well as self-reported diagnoses information (1). They also provided blood and saliva samples for genome-wide genotyping (see below). Several waves of follow-up assessments were conducted after initial assessment, including physical and mental health measurements in the form of mental health questionnaire (2).

This project has received approval to be conducted using the UK Biobank resource, under the project application number 82087.

##### Genotyping

All participants in UKB underwent genome-wide genotyping by either the UKB Axiom or the UK BiLEVE Axiom arrays, identifying 805,426 single nucleotide polymorphisms (SNPs) for 488,377 UKB participants (1). SNPs present on both genotyping assays above underwent quality control (QC), filtering SNPs with genotype missing calls > 5%, minor allele frequencies (MAF) < 0.0001 and failing QCs in multiple batches (1). This resulted in 670,739 SNPs being imputed using SHAPEIT3 in 487,442 participants, after filtering for missing genotype calls and heterozygosity in samples (1). Haplotype imputation was performed using Haplotype Reference Consortium (HRC) (3) and the merged UK10K and 1000 Genomes Phase 3 reference panels (4). This eventually resulted in 93,095,623 autosomal SNPs for UKB participants passing quality control steps (1).

Further QC was performed on genotyped data provided by the UKB. Genotyped SNPs were filtered off with missing genotyping rates (GENO) > 0.02, Hardy-Weinberg Equilibrium (HWE) p-value < 10^-8^ and minor allele frequencies (MAF) < 0.01. Individuals were filtered for abnormal heterozygosity (as defined during centralised quality control). Population structure was assessed using 4-means clustering in the first two principal components (PCs), and only participants of European ancestry were retained. Relatedness of individuals was handled by REGENIE in subsequent analyses (5). Imputed SNPs were further filtered with an imputation quality (INFO) score threshold of 0.4.

##### Primary care records

According to documentation from UKB, there is currently no national system for collecting primary care records (6). Therefore, at the current release, UKB liaised with data suppliers and other intermediaries to establish linkage to primary care data for ~230,000 (~ 45%) UKB participants, all of whom have provided written consent for linkage to their health-related records.

For primary care records, clinical events were coded by READ v2 and CTV-3 (6). Prescription records in the primary care setting were coded by READ v2, British National Formulary (BNF) and Dictionary of Medicines and Devices (dm+d) (6). Prescriptions are available from the 1990s to 2018, with the start and end dates dependent on the databases linked to specific regions of practice. Only the dates of prescription are available with no information on dispensing. Prescription codes for SSRIs were extracted from all available records. Further details of coding systems, including READ codes, BNF and dm+d codes are available in documentations from UKB (6) and previous work on treatment-resistant depression (7). Details of the data providers and the coding schema used are summarised below.

| **Country** | **GP Computer System Supplier** | **Approx no. of UK Biobank participants** | **Clinical coding classification** | **Prescription coding** |
| --- | --- | --- | --- | --- |
| Scotland ^a^ | EMIS ^b^ / Vision ^c^ | 27,000 | - Read v2 | - Read v2 - British National Formulary (BNF) |
| Wales ^d^ | EMIS / Vision | 21,000 | - Read v2 | - Read v2 |
| England | TPP^e^ | 165,000 | - Clinical Terms Version 3 (CTV3 or Read v3) | - BNF |
|  | Vision | 18,000 | - Read v2 | - Read v2 - Dictionary of - Medicines and Devices (dm+d) |

^a^ UK Biobank has engaged Albasoft (http://www.albasoft.co.uk/) (a third party data processor) to obtain data from GP practices in Scotland.

^b^ EMIS Health (https://www.emishealth.com/) is a computer system supplier to the NHS and provides the EMIS Web practice management system.

^c^ Vision Health (https://www.visionhealth.co.uk/) (previously InPS) is a computer system supplier and provides the Vision practice management system.

^d^ Data from Wales have been obtained via the SAIL Databank (https://saildatabank.com/) hosted by the University of Swansea.

^e^ TPP (https://www.tpp-uk.com/) is a computer system supplier and provides the SystmOne practice management system.

#### Replication Sample – Generation Scotland

##### Sample description

Generation Scotland (GenScot) is a family-based longitudinal study consisting around 24,000 volunteers to study the genetic and demographic characteristics of non-communicable disorders in Scotland (8). Participants aged between 35 and 65 years (and at least one first-degree relative aged above 18) were selected from primary care centres in Scotland from 2006 to 2011, resulting in a cohort of 23,960 participants. Participants provided blood and saliva samples for processing, including genome-wide genotyping. Notably, participants consented to the use of their linked electronic health records for research purposes. Linked data available for the GenScot cohort includes community-dispensed medications (Prescribing Information System, provided by Public Health Scotland), GP records (provided by Albasoft), inpatient, outpatient and registry records (Scottish Morbidity Records, provided by Public Health Scotland), and national death records (provided by Public Health Scotland). A depression-focussed follow-up study was conducted for between 2015 and 2017, consisting of remote mental health questionnaires (N = 9,618) and in-person clinic visits (N = 1,188) (9).

#### Polygenic scoring

Polygenic scores (PGS) for major depressive disorder (MDD), antidepressant non-remission and schizophrenia were computed using PRS-CS-auto (10) under the GenoPred pipeline (version 1, 7-Feb-2024) (11). The study characteristics and details of GWAS summary statistics are available in **Supplementary table 3**. All three GWAS (12–14) were performed by the Psychiatric Genomics Consortium, with no UKB samples included in the meta-analysis. The technical details for GenoPred (11) pipeline is available at: <https://opain.github.io/GenoPred/pipeline_technical.html>.

To handle relatedness issues in association testing between switching and PGS, only one of the two related pairs with kinship coefficient < 0.044 (third-degree relatives) was retained (See also **Methods** in main text). Kinship coefficients were estimated using the KING (15) software, as detailed in UKB documentation (1).

GWAS summary statistics underwent quality control as per documentation in GenoPred pipeline, filtering SNPs with duplicated rsids, INFO scores < 0.9, MAF < 0.01, and invalid p-values (p < 0 or p > 1). Variants in GWAS summary statistics were also matched to a reference panel consisting of HapMap3 variants (16) based on 1000 Genomes Phase 3 (1KG) and Human Genome Diversity Project (HGDP) samples. Strand-flipping was performed based on matching with this genotype reference, and SNPs with MAF differences < 0.2 between GWAS summary statistics and reference panel were removed.

PRS-CS is a polygenic scoring method that performs Bayesian shrinkage on the effect size distribution of variants using a continuous shrinkage prior (10). The “auto” framework under PRS-CS utilised a half-Cauchy prior on the phi (Φ) hyperparameter which controlled the degree of sparseness of the distribution of SNP effect sizes (10), such that Φ was estimated from target data, without the need for an external validation dataset. In GenoPred pipeline, PGS were computed in UKB, and reference PGS were also computed under the same framework consisting of 1KG European samples (17). The PGS computed in target samples were then re-scaled such that the mean and standard deviations were comparable to the 1KG European reference, so that the distributions of PGS and the results were comparable across samples.

#### Genome-wide association studies (GWASs)

Two GWASs were performed on patients with at least one and two GP diagnoses for MDD respectively using REGENIE (5). REGENIE divides association testing into 2 steps to enhance computational efficiency, and is well-suited to conduct genetic analyses on UKB. In Step 1, ridge regression was fitted into genome-wide genotyped SNPs divided into consecutive blocks of 1000 SNPs. The blocks of predictors from ridge regression were combined by another logistic ridge regression, with shrinkage parameters cross-validated by the leave-one-chromosome-out (LOCO) approach. These LOCO predictors were then used as covariates in Step 2 of REGENIE. Before running step 1 of REGENIE, the genotyped SNPs were pruned by linkage disequilibrium (LD), using R^2^ threshold of 0.9 within a window size of 1000 markers and a block size of 100 markers (5). In Step 2, association testing was performed on imputed SNPs genome-wide (filtered by INFO < 0.4) and switching status in both samples, adjusting for LOCO predictors and additional covariates (see below). In both steps of REGENIE, associations were further adjusted for genetic sex, index date of SSRI and first ten PCs for population stratification. Firth correction was used to correct for inflated SNP effect sizes and subsequent Type 1 error, which was evident when sample prevalence figures were low (5).

#### SNP-based heritability (h^2^_SNP_) estimation

h^2^_SNP_ estimation was performed on individual-level genotype data of both samples using two different methods, including genome-wide complex trait analysis (GCTA) version 1.94.1 (18) and Genome-wide Complex Trait Bayesian (GCTB) (19). Due to a lack of information on population prevalence of switching from SSRIs, h^2^_SNP_ estimates were reported on an observed scale.

GCTA estimates SNP-based heritability under the genomic relatedness–based restricted maximum likelihood (GREML) framework (18). Genomic relationship matrices (GRM) were constructed using genotyped SNPs that passed QC steps (see **Genotyping**) and further adjusted for incomplete tagging of causal SNPs. Genetic variance is subsequently estimated by fitting effects of genotyped SNPs (as GRM) in mixed linear model by REML. Related subjects were removed from the GRM with a cut-off at 0.05, and adjusted for covariates including first ten PCs for population stratification, genetic sex and index date of SSRI.

We also used the GCTB Bayes S framework in h^2^_SNP_ estimation under the default settings of GCTB (19,20), using 21000 iterations in Markov chain Monte Carlo (MCMC) with the first 1000 MCMC chains discarded / burnt in. GCTB framework allows simultaneous estimation SNP effects and genetic architecture characteristics, such as degrees of polygenicity (20). Therefore in GCTB, the degree of polygenicity (Pi) and negative selection (S) were also reported. By default, GCTB constrained h^2^_SNP_ to between 0 and 1 when running iterations for h^2^_SNP_ estimation. As a result, h^2^_SNP_ may be skewed and not normally distributed if true h^2^_SNP_ is close to zero. To support the use of posterior mean and standard errors in h^2^_SNP_ estimation, we reported the distributions of h^2^_SNP_ estimates across all iterations, and with 1000 burn-in iterations removed in **Supplementary figure 13** support the distributions are normally distributed. To ensure the robustness of our findings, we further reported the posterior mode and the 95% highest posterior density (HPD) intervals of h^2^_SNP_ estimates using GCTB, computed using the R package “HDInterval” (21).

### Supplementary tables

#### Supplementary table 1. READ v3 diagnosis codes used in this study

| Diagnosis | READ v3 code list |
| --- | --- |
| Depression | XE1YC, XE1ZZ, E1120, E1123, XaX53, E1124, E112z, E1122, E1121, E1125, E1126, XE1Y0, XSGom, XSGon, XE1ZY, X761L, E1130, E1133, XaX54, E1134, E1132, E1131, E1135, E1136, XE1Y1, E113z, XE1Zf, Eu334, XE1Zd, XE1Ze, Eu331, Eu330, XE1Zc, E1137, E291., Eu33y, XE1Za, XSGol, Eu321, X00Sb, XSGok, Eu320, XaCIs, X00SU, E2112, XE1Zb, E2B.., E2B1., E290z, E11y2, XaY2C, X00SQ, E112., E113., E11z2, E135., E2003, E204., Eu322, Eu323, Eu324, Eu325, Eu326, Eu32y, Eu32z, Eu33., Eu332, Eu333, Eu33z, Eu34., Eu3y1, Eu412, X00SO, X00SR, X00SS, Xa0wV, XaB9J, XaCHr, XaCHs, XaCIu, XE1aY, XM1GC, XSEGJ, XE1Xy |
| Bipolar disorders | XE1Xz, E11y0, E1170, E1173, E1174, E117z, E1172, E1171, E1175, E1176, E117., E1100, E1103, E1104, E1102, E1101, E1105, E1106, E1110, E1113, E1114, E1112, E1111, E1115, E1116, E111., E111z, E11z1, E11y3, Eu30y, XE1ZX, E11yz, E11y., E1160, E1163, E1164, E116z, E1162, E1161, E1165, E1166, E116., XE1ZW, E110z, Eu301, XE1ZV, X00SL, E2113, X00SN, XaY1Y, X00SM, Eu31z, E1140, E1143, E1144, E114z, E1142, E1141, E1145, E1146, Eu317, E1150, E1153, E1154, E115z, E1152, E1151, E1155, E1156, Eu314, Eu316, Eu313, Eu311, Eu312, E114., Eu310, E115., E11y1, E11.., E110., E11y2, E11z., E11z0, E2111, Eu30., Eu300, Eu302, Eu30z, Eu31., Eu315, Eu31y |
| Psychosis | XE1Xw, XE1Xy, XE1ZQ, Xa0s9, XE1ZR, XE2b8, E1000, E1070, E03y3, E1030, XE1ZU, E1010, E1020, Eu203, E1001, E1071, E1031, E1011, E1021, E100z, E100., E10.., Eu20z, Eu2.., E10z., E1005, Eu20., E107z, E1075, E107., Eu25z, XE2uT, XE2un, Eu25., E106., E130., E134., X00S8, E103z, E1035, E103., E12z., E1y.., E10yz, XE1Xx, XE1ZM, Eu25y, E13yz, E13y., XE1ZT, XE1Y3, Eu23y, XE1Y5, XaX52, E1..., E101z, E1015, E101., E1002, E1072, E1032, XE1Y2, E1012, E1022, E10y1, E102z, E1025, E102., E13y1, E10y0, X00SC, Eu231, E131., E1003, E1073, E1033, E1013, E1023, E1004, E1074, E1034, E1014, E1024, XE1ZS, E104., E10y., E11zz, E121., E123., E13.., E13z., E1410, E1411, E141z, Eu0z., Eu200, Eu201, Eu202, Eu204, Eu205, Eu206, Eu20y, Eu220, Eu230, Eu232, Eu233, Eu23z, Eu252, Eu2y., Eu2z., X00S6, X761M, XE1aM, XE1aO |
| Substance abuse | XE1Xu, XE1YR, XE1YS, XE1YY, XE1YZ, XE1Ya, XE1ZH, XE1ZI, XE1ZJ, XE2um, XE2n9, E0112, E2400, E2310, Xa1bX, E022., E014., E2470, E247z, E2473, E2472, E2471, XE1YW, E02yz, E02y., E01yz, E01y., E240z, E2403, E25y0, E25yz, E25y3, E25y2, E25y1, XE1Yb, E2550, E255z, E2553, E2552, E2551, E255., E2590, E259z, E2593, E2592, E2591, E259., E2540, E254z, E2543, E2542, E2541, E2530, E253z, E2533, E2532, E2531, E2560, E256z, E2563, E2562, E2561, E256., E2520, E252z, E2523, E2522, E2521, E252., E2580, E258z, E2583, E2582, E2581, E2570, E257z, E2573, E2572, E2571, E2500, E250z, E2503, E2502, E2501, XE1YX, E2594, E25z., Eu184, Eu183, Eu18z, Eu187, Eu185, Eu18y, Eu181, Eu186, Eu180, Eu18., Eu174, Eu173, Eu17z, Eu177, Eu175, Eu17y, Eu171, Eu172, Eu176, Eu170, Eu134, Eu133, Eu13z, Eu137, Eu135, Eu13y, Eu131, Eu132, Eu136, Eu130, Eu13., Eu154, Eu153, Eu15z, Eu157, Eu155, Eu15y, Eu151, Eu156, Eu150, Eu15., Eu114, Eu11z, Eu117, Eu115, Eu11y, Eu111, Eu116, Eu110, Eu11., Eu164, Eu163, Eu16z, XE1ZK, Eu165, Eu16y, Eu161, Eu166, Eu160, Eu16., XaKC3, XaKC2, XaKC8, XaKC6, XaKC4, XaKC7, XaKC0, XaKC1, XaKC5, XaKBz, XaKBy, Eu144, Eu143, Eu14z, Eu147, Eu145, Eu14y, Eu141, Eu142, Eu146, Eu140, Eu14., Eu124, Eu123, Eu12z, Eu127, Eu125, Eu12y, Eu121, Eu122, Eu126, Eu120, Eu12., Eu104, Eu103, Eu10z, XE1ZG, XE1ZF, Eu10y, Eu101, XE1ZE, Eu106, Eu100, Eu10., Eu1.., Eu194, Eu193, Eu19z, Eu197, Eu195, Eu19y, Eu191, XE1ZL, Eu196, Eu190, Eu19., Eu17., E0111, E011., E2410, E241z, E2413, E2412, E2411, E2450, E245z, E2453, E2452, E2451, XE1YV, E2460, E246z, E2463, E2462, E2461, E246., E2402, E2312, E2302, E24A., E020., E02z., E02.., E02y4, E0210, E021., E021z, E0211, E02y3, E02y1, E02y0, E02y2, E24z., E24.., E010., E2401, E2311, E2301, E2480, E248z, E2483, E2482, E2481, E248., E249., E2490, E249z, E2493, E2492, E2491, E242., E242z, E2420, E2423, E2422, E2421, E231z, E2313, XE1YQ, E0120, XE1YT, E243z, E2430, E2433, E2432, E2431, E2440, E244z, E2443, E2442, E2441, XE1YU, E01z., E01.., E015., E01y0, XaLWu, E013., E23z., E011z, E2300, E2303, E230z, E230., E258., E0110, E012., E23.., E231., E240., E241., E243., E244., E245., E247., E250., E253., E254., E257., E25y., Eu102, Eu105, Eu107, Eu112, Eu113, Eu152, Eu162, Eu167, Eu182, Eu192, XE1b0, XE1b2, XE1b6 |
| Anxiety disorders | X7627, XaIvf, Xa3WJ, E200., E2000, XE1Y7, E2002, E2004, E2005, E200z, XE1YA, E2020, E2021, E2022, E2023, E2024, E2025, E2026, E2027, E2028, E2029, X762G, Xa7lj, E202C, X762T, E202E, XE1YB, Eu40., X00SV, X00SW, X00SX, X00SY, Eu40y, Eu40z, Eu41., Eu410, Eu413, XE1Zj, Eu41z, 146G., E2001, E202., E202A, E202B, E202D, E202z, E28z., Eu400, Eu401, Eu402, Eu403, Eu411, Eu41y, Ua1qa, Ua1qc, Ua1qd, Ua1qe, Ua1qS, Ua1qs, Ua1qt, Ua1qU, Ua1qV, Ua1qW, Ua1qX, Ua1qY, X00Sr, X00Sy, X50G2, X50G3, X50G5, X50G6, X50GI, X761a, X761b, X761c, X761d, X761e, X761f, X761G, X761g, X761h, X761i, X761j, X761k, X761l, X761m, X761n, X761p, X761q, X761r, X761R, X761t, X761T, X761U, X761u, X761v, X761V, X761w, X761W, X761x, X761X, X761y, X761Y, X761z, X761Z, X7620, X7621, X7622, X7623, X7624, X7625, X7626, X7628, X7629, X762a, X762A, X762b, X762B, X762C, X762E, X762F, X762H, X762I, X762J, X762K, X762L, X762M, X762N, X762O, X762P, X762R, X762S, X762U, X762V, X762W, X762X, X762Y, X762Z, Xa00r, Xa00s, Xa1a8, Xa1Ev, Xa3Vk, Xa3Vl, Xa3WI, XaB96, XaEKL, XaK2c, XE1aW |

#### Supplementary table 2. Antidepressant names and respective drug classes

| Antidepressant name | Antidepressant drug Class |
| --- | --- |
| Sertraline | Selective serotonin reuptake inhibitors |
| Citalopram | Selective serotonin reuptake inhibitors |
| Fluoxetine | Selective serotonin reuptake inhibitors |
| Escitalopram | Selective serotonin reuptake inhibitors |
| Paroxetine | Selective serotonin reuptake inhibitors |
| Fluvoxamine | Selective serotonin reuptake inhibitors |
| Dapoxetine | Selective serotonin reuptake inhibitors |
| Moclobemide | Monoamine oxidase inhibitors |
| Phenelzine | Monoamine oxidase inhibitors |
| Tranylcypromine | Monoamine oxidase inhibitors |
| Isocarboxazid | Monoamine oxidase inhibitors |
| Tranylcypromine_trifluoperazine | Combination |
| Bupropion | Norepinephrine and dopamine reuptake inhibitors |
| Reboxetine | Norepinephrine reuptake inhibitors |
| Viloxazine | Norepinephrine reuptake inhibitors |
| Trazodone | Serotonin antagonist and reuptake inhibitors |
| Nefazodone | Serotonin antagonist and reuptake inhibitors |
| Venlafaxine | Serotonin-norepinephrine reuptake inhibitors |
| Duloxetine | Serotonin-norepinephrine reuptake inhibitors |
| Mirtazapine | Other antidepressants |
| Vortioxetine | Other antidepressants |
| Agomelatine | Other antidepressants |
| Flupentixol | Phenothiazine antipsychotics |
| Mianserin | Tetracyclic antidepressants |
| Maprotiline | Tetracyclic antidepressants |
| Amitriptyline | Tricyclic antidepressants |
| Imipramine | Tricyclic antidepressants |
| Dosulepin | Tricyclic antidepressants |
| Lofepramine | Tricyclic antidepressants |
| Nortriptyline | Tricyclic antidepressants |
| Clomipramine | Tricyclic antidepressants |
| Trimipramine | Tricyclic antidepressants |
| Doxepin | Tricyclic antidepressants |
| Amoxapine | Tricyclic antidepressants |
| Protriptyline | Tricyclic antidepressants |
| Amitriptyline_perphenazine | Combination |
| Nortriptyline_fluphenazine | Combination |

**Caption**

Antidepressant names are active ingredients listed as strings in primary care records in UK Biobank.

#### Supplementary table 3. GWAS summary statistics used for polygenic scoring

| Phenotype | Author (s) | Publication Year | Ancestry | Sample size | Number of SNPs |
| --- | --- | --- | --- | --- | --- |
| Non-remission | Pain et al. (12) | 2022 | European | 5,151 Remission 1,852  Non-remission 3,299 | 9,612,897 |
| MDD | Wray et al. (13) | 2018 | European | 143,265  Cases 45,591  Controls 97,674 | 11,345,536 |
| Schizophrenia | Trubetskoy et al. (14) | 2022 | European | 130,644 Cases 53,386  Controls 77,258 | 7,659,767 |

**Abbreviations**

MDD = major depressive disorder.

#### Supplementary table 4. Number of antidepressant prescriptions in primary sample, UK Biobank (by drug class and diagnosis)

| Sample | Full Sample (N = 71,153*^1^*) | ≥ 1 depression diagnosis record (N = 43,510*^1^*) | ≥ 2 depression diagnosis records (N = 29,687*^1^*) |
| --- | --- | --- | --- |
| **Drug class** |  |  |  |
| Combination | 2 (1, 6) | 3 (1, 6) | 2 (1, 6) |
| MAOI | 7 (2, 25) | 6 (2, 25) | 6 (2, 26) |
| NDRI | 2 (1, 2) | 1 (1, 2) | 1 (1, 2) |
| NRI | 3 (1, 10) | 3 (1, 9) | 4 (1, 9) |
| Other antidepressants | 5 (1, 17) | 4 (1, 16) | 4 (1, 16) |
| SARI | 4 (1, 16) | 4 (1, 14) | 4 (1, 15) |
| SNRI | 8 (2, 30) | 9 (2, 32) | 10 (3, 34) |
| SSRI | 18 (7, 47) | 22 (9, 54) | 25 (10, 60) |
| TCA | 6 (2, 22) | 6 (2, 22) | 6 (2, 23) |
| Tetracyclics | 3 (1, 9) | 4 (2, 8) | 4 (2, 8) |

**Caption**

*^1^* Number of prescriptions: Median (IQR).

**Abbreviations**

IQR = interquartile range; MAOI = monoamine oxidase inhibitors; NDRI = norepinephrine-dopamine reuptake inhibitors; NRI = norepinephrine reuptake inhibitors; SARI = serotonin antagonist and reuptake inhibitors; SNRI = serotonin–norepinephrine reuptake inhibitors; SSRI = selective serotonin reuptake inhibitors; TCA = tricyclic antidepressants.

#### Supplementary table 5. Number of patients with at least one prescription for antidepressant drug classes in primary sample, UK Biobank (by diagnosis)

| Sample | Full Sample (N = 38,813^1^) | ≥ 1 depression diagnosis record (N = 22,706*^1^*) | ≥ 2 depression diagnosis records (N = 14,781*^1^*) |
| --- | --- | --- | --- |
| **Drug class** |  |  |  |
| SSRI | 38813 (100%) | 22706 (100%) | 14781 (100%) |
| TCA | 18125 (46.7%) | 11320 (49.9%) | 7749 (52.4%) |
| Other antidepressants | 5268 (13.6%) | 3517 (15.5%) | 2623 (17.7%) |
| SNRI | 5139 (13.2%) | 3471 (15.3%) | 2624 (17.8%) |
| SARI | 2285 (5.9%) | 1498 (6.6%) | 1126 (7.6%) |
| Combination | 831 (2.1%) | 552 (2.4%) | 409 (2.8%) |
| NRI | 284 (0.7%) | 187 (0.8%) | 160 (1.1%) |
| NDRI | 189 (0.5%) | 105 (0.5%) | 85 (0.6%) |
| MAOI | 165 (0.4%) | 117 (0.5%) | 99 (0.7%) |
| Tetracyclics | 54 (0.1%) | 37 (0.2%) | 31 (0.2%) |

**Caption**

*^1^* Number of patients: count (percentage).

**Abbreviations**

MAOI = monoamine oxidase inhibitors; NDRI = norepinephrine-dopamine reuptake inhibitors; NRI = norepinephrine reuptake inhibitors; SARI = serotonin antagonist and reuptake inhibitors; SNRI = serotonin–norepinephrine reuptake inhibitors; SSRI = selective serotonin reuptake inhibitors; TCA = tricyclic antidepressants.

#### Supplementary table 6. Number of patients with depression and anxiety diagnoses on or before index SSRI exposure in primary sample, UK Biobank (by diagnosis)

| **Sample** | **Sample size** | **Proportion of patients with GP diagnosis on or before SSRI exposure** | |
| --- | --- | --- | --- |
|  |  | **Depression** | **Depression or anxiety** |
| ≥ 1 depression diagnosis record | 22706 | 16962 (74.7%) ^a^ | 17525 (77.2%) |
| ≥ 2 depression diagnoses records | 14781 | 11502 (77.8%) | 11902 (80.5%) |

^a^ Dates of depression diagnosis was not available for 39 participants in this sample.

**Abbreviations**

SSRI = selective serotonin reuptake inhibitors.

#### Supplementary table 7. Summary for Generation Scotland replication sample

| Characteristic | Non-switchers (N = 1,279*^1^*) | Switchers (N = 498*^1^*) | p-value*^2^* |
| --- | --- | --- | --- |
| Index SSRI |  |  | 0.017 |
| Fluoxetine | 433 (34%) | 165 (33%) |  |
| Citalopram | 604 (47%) | 190 (38%) |  |
| Sertraline | 199 (16%) | 125 (25%) |  |
| Escitalopram | 19 (1.5%) | 11 (2.2%) |  |
| Paroxetine | 24 (1.9%) | 7 (1.4%) |  |
| Age on index date | 47 (36, 56) | 51 (39, 62) | <0.001 |
| Unknown | 9 | 0 |  |
| Time to first switch | - | 31 (31, 61) |  |
| Sex |  |  | 0.10 |
| Female | 989 (77%) | 366 (73%) |  |
| Male | 290 (23%) | 132 (27%) |  |
| Ethnic background |  |  | 0.019 |
| White | 1,179 (98%) | 450 (97%) |  |
| Asian | 1 (<0.1%) | 5 (1.1%) |  |
| Mixed | 3 (0.3%) | 3 (0.6%) |  |
| Other | 2 (0.2%) | 0 (0%) |  |
| Not specified | 13 (1.1%) | 8 (1.7%) |  |
| Unknown | 81 | 32 |  |
| Body mass index | 26.2 (23.1, 30.2) | 25.8 (22.3, 29.5) | 0.057 |
| Unknown | 68 | 31 |  |
| Neuroticism score | 6 (4, 9) | 6 (3, 9) | 0.10 |
| Unknown | 173 | 60 |  |
| Family history of depression | 335 (27%) | 123 (26%) | 0.7 |
| Unknown | 32 | 19 |  |
| Qualifications |  |  | 0.034 |
| None of the above | 76 (6.7%) | 37 (8.9%) |  |
| Secondary | 264 (23%) | 116 (28%) |  |
| Vocational | 253 (22%) | 93 (22%) |  |
| Further | 134 (12%) | 44 (11%) |  |
| University Degree | 331 (29%) | 91 (22%) |  |
| Not available | 81 (7.1%) | 36 (8.6%) |  |
| Unknown | 140 | 81 |  |
| Annual income |  |  | 0.7 |
| Less than 10,000 | 121 (11%) | 55 (14%) |  |
| 10,000-30,000 | 374 (34%) | 141 (35%) |  |
| 30,000-50,000 | 280 (25%) | 106 (26%) |  |
| 50,000-70,000 | 164 (15%) | 41 (10%) |  |
| Greater than 70,000 | 81 (7.4%) | 21 (5.2%) |  |
| Not available | 82 (7.4%) | 42 (10%) |  |
| Scottish Index of Multiple Deprivation Quintile |  |  | 0.5 |
| 1 | 209 (17%) | 93 (20%) |  |
| 2 | 200 (17%) | 81 (17%) |  |
| 3 | 213 (18%) | 75 (16%) |  |
| 4 | 281 (23%) | 96 (21%) |  |
| 5 | 304 (25%) | 122 (26%) |  |
| Unknown | 72 | 31 |  |

**Caption**

^1^ n (%); Median (IQR).

^2^ Kruskal-Wallis rank sum test; Wilcoxon rank sum test; Pearson’s Chi-squared test.

**Abbreviations**

SSRI = selective serotonin reuptake inhibitors

#### Supplementary table 8. Switching patterns in primary sample, UK Biobank

| **SSRI (pre-switch)** | |  | **Fluoxetine (**N = 1,687^1^) | **Paroxetine (**N = 621^1^) | **Citalopram  (**N = 1,586^1^) | **Sertraline**  (N = 1,024^1^) | **Escitalopram**  (N = 201^1^) | **Fluvoxamine**  (N = 14^1^) | **Overall**  (N = 5133^1^) |
| --- | --- | --- | --- | --- | --- | --- | --- | --- | --- |
| **Drug class (post-switch)** | **TCA** |  | 572 (34%) | 248 (40%) | 467 (29%) | 254 (25%) | 49 (24%) | 7 (50%) | **1597 (31%)** |
|  | **NRI** |  | 3 (0.2%) | 4 (0.6%) | 4 (0.3%) | 2 (0.2%) | 0 (0%) | 0 (0%) | **13 (0.3%)** |
|  | **Another SSRI** |  | 785 (47%) | 273 (44%) | 737 (46%) | 478 (47%) | 102 (51%) | 5 (36%) | **2380 (46%)** |
|  | **Combination** |  | 14 (0.8%) | 14 (2.3%) | 16 (1.0%) | 7 (0.7%) | 4 (2.0%) | 0 (0%) | **55 (1.1%)** |
|  | **Other antidepressants** |  | 131 (7.8%) | 14 (2.3%) | 199 (13%) | 172 (17%) | 17 (8.5%) | 1 (7.1%) | **534 (10%)** |
|  | **SNRI** |  | 118 (7.0%) | 49 (7.9%) | 111 (7.0%) | 79 (7.7%) | 24 (12%) | 0 (0%) | **381 (7.4%)** |
|  | **SARI** |  | 62 (3.7%) | 15 (2.4%) | 50 (3.2%) | 30 (2.9%) | 5 (2.5%) | 1 (7.1%) | **163 (3.2%)** |
|  | **MAOI** |  | 1 (<0.1%) | 2 (0.3%) | 2 (0.1%) | 2 (0.2%) | 0 (0%) | 0 (0%) | **7 (0.1%)** |
|  | **NDRI** |  | 0 (0%) | 2 (0.3%) | 0 (0%) | 0 (0%) | 0 (0%) | 0 (0%) | **2 (< 0.1%)** |
|  | **Tetracyclics** |  | 1 (<0.1%) | 0 (0%) | 0 (0%) | 0 (0%) | 0 (0%) | 0 (0%) | **1 (<0.1%)** |

**Caption**

^1^ Number of patients with SSRI pre-switching: count (percentage).

**Abbreviations**

MAOI = monoamine oxidase inhibitors; NDRI = norepinephrine-dopamine reuptake inhibitors; NRI = norepinephrine reuptake inhibitors; SARI = serotonin antagonist and reuptake inhibitors; SNRI = serotonin–norepinephrine reuptake inhibitors; SSRI = selective serotonin reuptake inhibitors; TCA = tricyclic antidepressants.

#### Supplementary table 9. Switching patterns in Generation Scotland replication sample

| **SSRI (pre-switch)** | |  | **Fluoxetine (**N = 165^1^) | **Paroxetine (**N = 7^1^) | **Citalopram  (**N = 190^1^) | **Sertraline**  (N = 125^1^) | **Escitalopram**  (N = 11^1^) | **Fluvoxamine**  0 (0%) | **Overall**  (N = 498^1^) |
| --- | --- | --- | --- | --- | --- | --- | --- | --- | --- |
| **Drug class (post-switch)** | **TCA** |  | 26 (15.8%) | 0 (0%) | 30 (15.8%) | 16 (12.8%) | 1 (9.1%) | 0 (0%) | **73 (14.7%)** |
|  | **NRI** |  | 0 (0%) | 0 (0%) | 0 (0%) | 0 (0%) | 0 (0%) | 0 (0%) | **0 (0%)** |
|  | **Another SSRI** |  | 81 (49.1%) | 2 (28.6%) | 99 (52.1%) | 48 (38.4%) | 7 (63.6%) | 0 (0%) | **237 (47.6%)** |
|  | **Combination** |  | 0 (0%) | 0 (0%) | 0 (0%) | 0 (0%) | 0 (0%) | 0 (0%) | **0 (0%)** |
|  | **Other antidepressants** |  | 37 (22.4%) | 2 (28.6%) | 42 (22.1%) | 45 (36.0%) | 2 (18.2%) | 0 (0%) | **128 (25.7%)** |
|  | **SNRI** |  | 17 (10.3%) | 3 (42.8%) | 16 (8.4%) | 13 (10.4%) | 1 (9.1%) | 0 (0%) | **50 (10.0%)** |
|  | **SARI** |  | 4 (2.4%) | 0 (0%) | 3 (1.6%) | 2 (1.6%) | 0 (0%) | 0 (0%) | **9 (1.8%)** |
|  | **MAOI** |  | 0 (0%) | 0 (0%) | 0 (0%) | 1 (0.8%) | 0 (0%) | 0 (0%) | **1 (0.2%)** |
|  | **NDRI** |  | 0 (0%) | 0 (0%) | 0 (0%) | 0 (0%) | 0 (0%) | 0 (0%) | **0 (0%)** |
|  | **Tetracyclics** |  | 0 (0%) | 0 (0%) | 0 (0%) | 0 (0%) | 0 (0%) | 0 (0%) | **0 (0%)** |

**Caption**

^1^ Number of patients with SSRI pre-switching: count (percentage).

**Abbreviations**

MAOI = monoamine oxidase inhibitors; NDRI = norepinephrine-dopamine reuptake inhibitors; NRI = norepinephrine reuptake inhibitors; SARI = serotonin antagonist and reuptake inhibitors; SNRI = serotonin–norepinephrine reuptake inhibitors; SSRI = selective serotonin reuptake inhibitors; TCA = tricyclic antidepressants.

#### Supplementary table 10. Statistical tests for distributions of SSRI switcher / non-switcher status across assessment centres and genotyping batches

| Sample | Covariate | df | p-value |
| --- | --- | --- | --- |
| Full sample | Genotype batch | 105 | 0.346067 |
|  | Assessment centre | 21 | **2.54E-08*** |
| ≥ 1 depression diagnosis record | Genotype batch | 105 | 0.117948 |
|  | Assessment centre | 20 | **7.92E-06*** |
| ≥ 2 MDD diagnosis records | Genotype batch | 105 | 0.292841 |
|  | Assessment centre | 20 | **0.024137*** |

**Caption**

Statistical significance assessed by Kruskal-Wallis test.

* p-value < 0.05.

#### Supplementary table 11. Odds ratio for association testing in primary sample, UK Biobank

| Study outcome | | All sample | | ≥ 1 depression diagnosis record | | ≥ 2 depression diagnosis records | |
| --- | --- | --- | --- | --- | --- | --- | --- |
|  |  | **OR (95% CI)^1^** | **p^2^** | **OR (95% CI)^1^** | **p^2^** | **OR (95% CI)^1^** | **p^2^** |
| Educational levels | **None of the above** | Reference | | | | | |
|  | **Secondary** | 0.82 [0.75-0.89] |  | 0.87 [0.78-0.97] |  | 0.88 [0.77-1.00] |  |
|  | **Vocational** | 0.86 [0.77-0.95] |  | 0.89 [0.78-1.02] |  | 0.88 [0.74-1.04] |  |
|  | **Further** | 0.78 [0.70-0.87] |  | 0.86 [0.74-0.99] |  | 0.77 [0.65-0.92] |  |
|  | **University Degree** | 0.73 [0.67-0.79] | **1.53E-10**** | 0.78 [0.69-0.87] | **0.001**** | 0.79 [0.69-0.91] | **0.013*** |
| Annual income | **Less than £18,000** | Reference | | | | | |
|  | **£18,000-£31,000** | 0.87 [0.80-0.95] |  | 0.89 [0.80-0.99] |  | 0.88 [0.77-1.01] |  |
|  | **£31,000-£52,000** | 0.77 [0.71-0.84] |  | 0.8 [0.71-0.89] |  | 0.78 [0.68-0.90] |  |
|  | **£52,000-£100,000** | 0.7 [0.62-0.77] |  | 0.73 [0.63-0.83] |  | 0.72 [0.61-0.85] |  |
|  | **Greater than £100,000** | 0.66 [0.53-0.83] | **6.79E-15**** | 0.61 [0.44-0.82] | **4.92E-07**** | 0.75 [0.52-1.06] | **1.43E-04**** |
| Townsend Deprivation Index | | 1.01 [1.00-1.02] | 0.085 | 1.01 [0.99-1.02] | 0.268 | 1.00 [0.99-1.02] | 0.722 |
| Family history^3^ | | 1.08 [1.00-1.16] | **0.048*** | 1.09 [0.99-1.19] | 0.084 | 1.10 [0.99-1.23] | 0.088 |
| PGS_MDD_ | | 1.02 [0.99-1.06] | 0.138 | 1.00 [0.96-1.05] | 0.848 | 0.99 [0.94-1.04] | 0.73 |
| PGS_non-rem_ | | 1.03 [1.00-1.07] | **0.029*** | 1.04 [1.00-1.08] | **0.031*** | 1.07 [1.02-1.12] | **0.007**** |
| PGS_SCZ_ | | 1.02 [0.99-1.05] | 0.212 | 1.00 [0.96-1.04] | 0.903 | 0.99 [0.95-1.04] | 0.737 |

**Abbreviations**

MDD = major depressive disorder; non-rem = non-remission from antidepressants; OR = odds ratio; PGS = polygenic score; SCZ = schizophrenia; SSRI = selective serotonin reuptake inhibitors.

^1^ Associations controlled for index date of SSRI, sex, assessment centre and 10 principal components for population stratification.

^2^ p-values tested by likelihood ratio test.

^3^ Family history of severe depression.

* p < 0.05.

** p < 0.00714 (Bonferroni correction *n* = 7).

#### Supplementary table 12. Estimates for SNP-based heritability

| Sample | Method | h^2^_observed_ | SE | p-value |
| --- | --- | --- | --- | --- |
| Full sample | **GCTA** | **0.0268** | **0.0158** | **0.038*** |
|  | **GCTB** | **0.0242** | **0.0095** | **0.005**** |
| ≥ 1 depression diagnosis record | **GCTA** | **0.0431** | **0.0268** | **0.048*** |
|  | **GCTB** | **0.0398** | **0.016** | **0.005**** |
| ≥ 2 depression diagnosis records | **GCTA** | 0.0216 | 0.0394 | 0.293 |
|  | **GCTB** | 0.035 | 0.025 | 0.08 |

* p-value < 0.05.

** p-value < 0.01.

**Abbreviations**

GCTA = genome-wide complex trait analysis; GCTB = Genome-wide Complex Trait Bayesian; h^2^_observed_ = SNP-based heritability (on observed scale); LDSC = linkage disequilibrium score regression; SE = standard error.

#### Supplementary table 13. Top 40 hit SNPs with p < 1e-5 for SSRI switching (full sample)

| rsid | chr | pos | EA | MAF | BETA | SE | p | Nearest Gene | dist | func | CADD |
| --- | --- | --- | --- | --- | --- | --- | --- | --- | --- | --- | --- |
| rs56206563 | 8 | 33851064 | T | 0.0159 | 0.46512 | 0.0864874 | 2.53E-07 | RP11-317N12.1:RP1-273G13.3 | 00:00 | ncRNA_intronic | 1.729 |
| rs72630918 | 8 | 33659446 | G | 0.0338 | 0.377248 | 0.0735669 | 7.23E-07 | RP11-317N12.1 | 0 | ncRNA_intronic | 1.333 |
| rs1127390 | 16 | 81010073 | T | 0.2296 | -0.133574 | 0.0273119 | 8.20E-07 | CMC2 | 0 | exonic | 0.248 |
| rs9931292 | 16 | 81009568 | G | 0.2296 | -0.133457 | 0.0273087 | 8.36E-07 | CMC2 | 129 | downstream | 1.266 |
| rs10110388 | 8 | 33569261 | T | 0.02286 | -0.424367 | 0.0830298 | 8.78E-07 | RP11-317N12.1 | 0 | ncRNA_intronic | 0.194 |
| rs72630978 | 8 | 33786522 | A | 0.0159 | 0.455454 | 0.0889054 | 8.90E-07 | RP11-317N12.1 | 0 | ncRNA_intronic | 2.78 |
| rs9921448 | 16 | 81009490 | G | 0.2296 | -0.133087 | 0.0273102 | 8.98E-07 | CMC2 | 207 | downstream | 0.686 |
| rs2028594 | 8 | 33564880 | G | 0.02187 | -0.428417 | 0.0841754 | 9.95E-07 | RP11-317N12.1 | 0 | ncRNA_intronic | 0.097 |
| rs2549855 | 16 | 81026852 | G | 0.2326 | 0.132147 | 0.0272665 | 1.03E-06 | CMC2 | 0 | intronic | 0.96 |
| rs11378897 | 8 | 33924941 | TA | 0.0159 | -0.46628 | 0.0916406 | 1.04E-06 | RP11-317N12.1 | 28414 | intergenic | 5.422 |
| rs2123627 | 16 | 81021816 | A | 0.2336 | 0.13206 | 0.027274 | 1.06E-06 | CMC2 | 0 | intronic | 1.728 |
| rs874054 | 16 | 81006142 | C | 0.2296 | -0.1319 | 0.0272964 | 1.11E-06 | CMC2 | 3555 | intergenic | 3.58 |
| rs874053 | 16 | 81006107 | G | 0.2296 | -0.131886 | 0.0272961 | 1.11E-06 | CMC2 | 3590 | intergenic | 1.322 |
| rs61846889 | 10 | 50970429 | A | 0.1252 | 0.159793 | 0.0323949 | 1.12E-06 | OGDHL | 3 | upstream | 20.5 |
| rs56297803 | 16 | 81006252 | AT | 0.2296 | -0.131617 | 0.0273023 | 1.18E-06 | CMC2 | 3445 | intergenic | 0.336 |
| rs2911149 | 16 | 81026309 | A | 0.2336 | 0.131376 | 0.0272683 | 1.20E-06 | CMC2 | 0 | intronic | 0.352 |
| rs2602446 | 16 | 81017915 | C | 0.2316 | 0.131524 | 0.0273056 | 1.20E-06 | CMC2 | 0 | intronic | 4.031 |
| rs2549858 | 16 | 81026994 | A | 0.2306 | 0.131228 | 0.027258 | 1.22E-06 | CMC2 | 0 | intronic | 2.822 |
| rs2549859 | 16 | 81027003 | G | 0.2306 | 0.131228 | 0.027258 | 1.22E-06 | CMC2 | 0 | intronic | 4.164 |
| rs2549856 | 16 | 81026927 | T | 0.2326 | 0.131216 | 0.0272585 | 1.22E-06 | CMC2 | 0 | intronic | 1.951 |
| rs2549857 | 16 | 81026958 | A | 0.2326 | 0.131216 | 0.0272585 | 1.22E-06 | CMC2 | 0 | intronic | 0.139 |
| rs34844332 | 16 | 81009520 | AT | 0.2316 | -0.132042 | 0.0274481 | 1.24E-06 | CMC2 | 177 | downstream | 0.127 |
| rs2167890 | 16 | 81021920 | C | 0.1769 | 0.144854 | 0.0302595 | 1.31E-06 | CMC2 | 0 | intronic | 0.162 |
| rs2549825 | 16 | 81018159 | T | 0.1769 | 0.144631 | 0.0302891 | 1.39E-06 | CMC2 | 0 | intronic | 0.302 |
| rs2167889 | 16 | 81021472 | A | 0.2316 | 0.130433 | 0.0272486 | 1.40E-06 | CMC2 | 0 | intronic | 3.353 |
| 16:81023707_CT_C | 16 | 81023707 | C | 0.2326 | 0.130374 | 0.0272401 | 1.41E-06 | CMC2 | 0 | intronic | 0.564 |
| rs35195025 | 16 | 81023304 | TATC | 0.2316 | 0.130322 | 0.0272501 | 1.43E-06 | CMC2 | 0 | intronic | 7.19 |
| rs2549839 | 16 | 81023235 | T | 0.2316 | 0.1302 | 0.0272425 | 1.46E-06 | CMC2 | 0 | intronic | 0.783 |
| rs2549844 | 16 | 81023861 | C | 0.2316 | 0.130012 | 0.0272089 | 1.47E-06 | CMC2 | 0 | intronic | 5.334 |
| rs2549822 | 16 | 81015560 | T | 0.1769 | 0.144057 | 0.0302687 | 1.51E-06 | CMC2 | 0 | intronic | 3.954 |
| rs1477381 | 16 | 81014700 | C | 0.2296 | 0.130154 | 0.0273036 | 1.55E-06 | CMC2 | 0 | intronic | 6.362 |
| rs2549834 | 16 | 81022544 | T | 0.1769 | 0.143713 | 0.0302653 | 1.59E-06 | CMC2 | 0 | intronic | 1.066 |
| rs2911148 | 16 | 81021894 | G | 0.2316 | 0.129669 | 0.027242 | 1.61E-06 | CMC2 | 0 | intronic | 0.584 |
| rs2549833 | 16 | 81022232 | C | 0.2316 | 0.129652 | 0.0272409 | 1.61E-06 | CMC2 | 0 | intronic | 1.661 |
| rs2549832 | 16 | 81020141 | G | 0.1769 | 0.14356 | 0.0302571 | 1.62E-06 | CMC2 | 0 | intronic | 0.735 |
| rs2602433 | 16 | 81029003 | A | 0.2336 | 0.129635 | 0.0272479 | 1.63E-06 | CMC2 | 0 | intronic | 2.795 |
| rs2549838 | 16 | 81023210 | T | 0.2316 | 0.129549 | 0.0272385 | 1.64E-06 | CMC2 | 0 | intronic | 4.09 |
| rs2549836 | 16 | 81023081 | C | 0.2316 | 0.129551 | 0.0272389 | 1.64E-06 | CMC2 | 0 | intronic | 2.37 |
| rs2549837 | 16 | 81023115 | A | 0.2316 | 0.129543 | 0.0272387 | 1.64E-06 | CMC2 | 0 | intronic | 11.46 |
| rs2316923 | 16 | 81024821 | G | 0.2316 | 0.129442 | 0.0272403 | 1.67E-06 | CMC2 | 0 | intronic | 0.74 |

**Caption**

Annotation of variants was performed by FUMA tool (<https://fuma.ctglab.nl/>).

**Abbreviations**

chr = chromosome; dist = distance to the nearest gene; EA = effect allele; func = functions annotated from ANNOVAR; MAF = minor allele frequency; MDD = major depressive disorder; pos = position on hg19; rsid = variant ID; SE = standard error; SSRI = selective serotonin reuptake inhibitors.

#### Supplementary table 14. Top 40 hit SNPs with p < 1e-5 for SSRI switching (≥ 1 MDD diagnosis record)

| rsid | chr | pos | EA | MAF | BETA | SE | p | Nearest Gene | dist | func | CADD |
| --- | --- | --- | --- | --- | --- | --- | --- | --- | --- | --- | --- |
| rs113737796 | 14 | 60845897 | C | 0.03877 | 0.351487 | 0.068506 | 6.78E-07 | C14orf39 | 17289 | intergenic | 0.906 |
| rs17097347 | 14 | 60817408 | T | 0.03777 | 0.351048 | 0.068594 | 7.20E-07 | CTD-2568P8.1 | 26782 | intergenic | 0.906 |
| 14:60854762_TA_T | 14 | 60854762 | T | 0.03777 | 0.348426 | 0.068639 | 8.80E-07 | C14orf39 | 8424 | intergenic | 5.085 |
| rs191325339 | 14 | 60889751 | C | 0.03777 | 0.343496 | 0.068264 | 1.09E-06 | C14orf39 | 0 | intronic | 0.896 |
| rs17097534 | 14 | 60961735 | T | 0.03777 | 0.345691 | 0.068743 | 1.11E-06 | C14orf39:SALL4P7 | 00:00 | ncRNA_exonic | 8.925 |
| 14:60901561_GTTATA_G | 14 | 60901561 | G | 0.03777 | 0.342844 | 0.069111 | 1.53E-06 | C14orf39 | 0 | intronic | 0.735 |
| rs75823917 | 14 | 60981509 | A | 0.03777 | 0.338341 | 0.068936 | 1.96E-06 | C14orf39 | 0 | intronic | 3.913 |
| rs12273649 | 11 | 24509974 | T | 0.06262 | 0.270512 | 0.055663 | 2.06E-06 | LUZP2 | 8541 | intergenic | 0.705 |
| rs149792878 | 14 | 49350551 | T | 0.0159 | -0.58276 | 0.131428 | 2.07E-06 | SNORD112 | 60098 | intergenic | 0.179 |
| rs77206066 | 17 | 7959125 | C | 0.04374 | 0.330666 | 0.067733 | 2.19E-06 | RP11-599B13.3 | 417 | downstream | 5.157 |
| rs79837728 | 14 | 60998509 | G | 0.03777 | 0.338002 | 0.06923 | 2.21E-06 | RP11-1042B17.3 | 0 | ncRNA_intronic | 9.689 |
| rs116052952 | 2 | 2.41E+08 | C | 0.02286 | 0.462469 | 0.094058 | 2.36E-06 | AC093802.1 | 28247 | intergenic | 5.749 |
| 14:60900965_TATA_T | 14 | 60900965 | T | 0.03181 | 0.366986 | 0.075345 | 2.44E-06 | C14orf39 | 0 | intronic | 0.775 |
| rs2125950 | 8 | 2928234 | G | 0.4036 | -0.14009 | 0.029636 | 2.48E-06 | CSMD1 | 0 | intronic | 3.327 |
| rs61949308 | 13 | 47238572 | C | 0.02187 | -0.54658 | 0.124029 | 2.66E-06 | LRCH1 | 0 | intronic | 0.007 |
| rs2125951 | 8 | 2928227 | G | 0.4036 | -0.13937 | 0.029636 | 2.79E-06 | CSMD1 | 0 | intronic | 2.344 |
| rs11878951 | 19 | 48792490 | C | 0.174 | 0.176505 | 0.037358 | 3.05E-06 | ZNF114 | 1624 | intergenic | 3.59 |
| rs140183378 | 4 | 43354672 | A | 0.02783 | 0.398119 | 0.08258 | 3.30E-06 | RP11-1E6.1 | 7054 | intergenic | 0.55 |
| rs202177809 | 4 | 43269936 | GC | 0.02783 | 0.4279 | 0.088743 | 3.35E-06 | RP11-395F4.1 | 32962 | intergenic | 0.994 |
| rs146536531 | 4 | 43358827 | A | 0.02783 | 0.396469 | 0.082554 | 3.59E-06 | RP11-1E6.1 | 11209 | intergenic | 0.97 |
| rs147130265 | 4 | 43357778 | T | 0.02783 | 0.395476 | 0.082565 | 3.80E-06 | RP11-1E6.1 | 10160 | intergenic | 2.617 |
| rs9651583 | 11 | 24507798 | C | 0.06262 | 0.261887 | 0.05543 | 3.87E-06 | LUZP2 | 10717 | intergenic | 2.834 |
| rs17457944 | 4 | 43356830 | T | 0.02783 | 0.395137 | 0.082567 | 3.87E-06 | RP11-1E6.1 | 9212 | intergenic | 0.331 |
| rs142825483 | 4 | 43356776 | T | 0.02783 | 0.395119 | 0.082567 | 3.87E-06 | RP11-1E6.1 | 9158 | intergenic | 11.26 |
| rs138323020 | 4 | 43356669 | A | 0.02783 | 0.395033 | 0.082569 | 3.90E-06 | RP11-1E6.1 | 9051 | intergenic | 2.884 |
| rs17536050 | 4 | 43355721 | C | 0.02783 | 0.394709 | 0.082568 | 3.96E-06 | RP11-1E6.1 | 8103 | intergenic | 2.559 |
| rs140355025 | 4 | 43355434 | A | 0.02783 | 0.394516 | 0.082571 | 4.01E-06 | RP11-1E6.1 | 7816 | intergenic | 2.468 |
| rs75824620 | 4 | 43359332 | A | 0.02783 | 0.395775 | 0.082849 | 4.02E-06 | RP11-1E6.1 | 11714 | intergenic | 7.517 |
| rs149873481 | 4 | 43354979 | A | 0.02783 | 0.39437 | 0.082571 | 4.04E-06 | RP11-1E6.1 | 7361 | intergenic | 1.013 |
| 4:43358173_CTA_C | 4 | 43358173 | C | 0.02783 | 0.395357 | 0.082788 | 4.04E-06 | RP11-1E6.1 | 10555 | intergenic | 0.919 |
| rs150723362 | 4 | 43354843 | C | 0.02783 | 0.39432 | 0.082572 | 4.05E-06 | RP11-1E6.1 | 7225 | intergenic | 1.758 |
| rs10456430 | 6 | 11763008 | A | 0.04274 | -0.32428 | 0.072744 | 4.09E-06 | ADTRP | 0 | intronic | 4.281 |
| rs146109701 | 4 | 43354470 | T | 0.02783 | 0.396238 | 0.083205 | 4.31E-06 | RP11-1E6.1 | 6852 | intergenic | 0.172 |
| rs142999246 | 4 | 43354458 | T | 0.02783 | 0.396196 | 0.083207 | 4.32E-06 | RP11-1E6.1 | 6840 | intergenic | 0.146 |
| rs189742614 | 4 | 43354459 | G | 0.02783 | 0.396196 | 0.083207 | 4.32E-06 | RP11-1E6.1 | 6841 | intergenic | 1.258 |
| rs140176922 | 4 | 43352617 | C | 0.02783 | 0.392782 | 0.082552 | 4.38E-06 | RP11-1E6.1 | 4999 | intergenic | 1.201 |
| rs144747495 | 4 | 43354012 | G | 0.02783 | 0.392761 | 0.082558 | 4.39E-06 | RP11-1E6.1 | 6394 | intergenic | 1.448 |
| rs138315122 | 4 | 43354240 | C | 0.02783 | 0.392754 | 0.08256 | 4.40E-06 | RP11-1E6.1 | 6622 | intergenic | 5.636 |
| rs138685627 | 4 | 43353958 | G | 0.02783 | 0.392719 | 0.082559 | 4.41E-06 | RP11-1E6.1 | 6340 | intergenic | 0.817 |
| rs150954018 | 4 | 43353407 | A | 0.02783 | 0.392551 | 0.082558 | 4.45E-06 | RP11-1E6.1 | 5789 | intergenic | 0.521 |

**Caption**

Annotation of variants was performed by FUMA tool (<https://fuma.ctglab.nl/>).

**Abbreviations**

chr = chromosome; dist = distance to the nearest gene; EA = effect allele; func = functions annotated from ANNOVAR; MAF = minor allele frequency; MDD = major depressive disorder; pos = position on hg19; rsid = variant ID; SE = standard error; SSRI = selective serotonin reuptake inhibitors.

#### Supplementary table 15. Top 40 hit SNPs with p < 1e-5 for SSRI switching (≥ 2 MDD diagnosis records)

| rsid | chr | pos | EA | MAF | BETA | SE | p | Nearest Gene | dist | func | CADD |
| --- | --- | --- | --- | --- | --- | --- | --- | --- | --- | --- | --- |
| rs7669265 | 4 | 76612386 | G | 0.04771 | -0.468464 | 0.084515 | 9.05E-08 | G3BP2 | 0 | intronic | 1.906 |
| rs6531785 | 4 | 76557406 | G | 0.04374 | -0.478522 | 0.0864935 | 9.64E-08 | CDKL2 | 1505 | intergenic | 5.689 |
| rs12518815 | 5 | 25130699 | A | 0.3907 | -0.186432 | 0.037506 | 5.99E-07 | RP11-549K20.1 | 0 | ncRNA_intronic | 1.255 |
| rs114149662 | 1 | 192934797 | C | 0.01392 | 0.692656 | 0.134315 | 9.50E-07 | LINC01032 | 17409 | intergenic | 0.95 |
| rs116052952 | 2 | 240752825 | C | 0.02286 | 0.564921 | 0.111459 | 1.22E-06 | AC093802.1 | 28247 | intergenic | 5.749 |
| rs75382569 | 10 | 8052328 | G | 0.09046 | 0.295131 | 0.0601542 | 1.67E-06 | TAF3 | 0 | intronic | 0.371 |
| rs145453945 | 6 | 116666178 | A | 0.1342 | 0.225461 | 0.0466309 | 1.94E-06 | DSE:RP1-93H18.1 | 00:00 | ncRNA_intronic | 0.735 |
| rs11753710 | 6 | 116671302 | G | 0.1342 | 0.225467 | 0.0466323 | 1.94E-06 | DSE:RP1-93H18.1 | 00:00 | ncRNA_intronic | 0.424 |
| rs113698755 | 6 | 116678687 | TC | 0.1352 | 0.225673 | 0.0468486 | 2.11E-06 | DSE:RP1-93H18.1 | 00:00 | ncRNA_intronic | 0.975 |
| rs112711055 | 15 | 68339776 | G | 0.04175 | -0.551114 | 0.123753 | 2.27E-06 | PIAS1 | 6740 | intergenic | 7.168 |
| rs149792878 | 14 | 49350551 | T | 0.0159 | -0.726296 | 0.16712 | 2.32E-06 | SNORD112 | 60098 | intergenic | 0.179 |
| rs145987863 | 1 | 192964414 | C | 0.01392 | 0.658682 | 0.132881 | 2.35E-06 | ZNF101P2 | 606 | upstream | 7.748 |
| 6:116672075_GA_G | 6 | 116672075 | G | 0.1342 | 0.224654 | 0.0470485 | 2.58E-06 | DSE:RP1-93H18.1 | 00:00 | ncRNA_intronic | 3.445 |
| rs17077864 | 6 | 116672939 | G | 0.1342 | 0.222368 | 0.0467968 | 2.88E-06 | DSE:RP1-93H18.1 | 00:00 | ncRNA_intronic | 4.704 |
| rs141713005 | 1 | 193002886 | A | 0.01292 | 0.676011 | 0.137814 | 2.99E-06 | UCHL5 | 0 | intronic | 4.447 |
| rs73548973 | 6 | 116651294 | C | 0.1342 | 0.219056 | 0.0463135 | 3.17E-06 | DSE:RP1-93H18.1 | 00:00 | ncRNA_intronic | 4.464 |
| rs79325251 | 12 | 15197630 | C | 0.0328 | 0.393047 | 0.0819221 | 3.41E-06 | RP11-508P1.2 | 38012 | intergenic | 0.824 |
| rs141588653 | 12 | 15213368 | A | 0.03181 | 0.392222 | 0.0818963 | 3.55E-06 | RERG | 47348 | intergenic | 1.662 |
| rs2089518 | 5 | 25358348 | T | 0.2992 | 0.185837 | 0.0404418 | 3.68E-06 | RP11-184E9.2 | 36892 | intergenic | 0.287 |
| rs59504977 | 6 | 116674329 | C | 0.1352 | 0.219914 | 0.0468233 | 3.73E-06 | DSE:RP1-93H18.1 | 00:00 | ncRNA_intronic | 16.16 |
| rs112851235 | 13 | 21567608 | T | 0.01292 | -1.08379 | 0.268922 | 4.17E-06 | LATS2 | 0 | intronic | 0.71 |
| rs146584554 | 12 | 15177708 | A | 0.03479 | 0.387927 | 0.0817136 | 4.34E-06 | RP11-508P1.2 | 18090 | intergenic | 0.579 |
| rs7738448 | 6 | 116676685 | A | 0.1352 | 0.218288 | 0.0468421 | 4.43E-06 | DSE:RP1-93H18.1 | 00:00 | ncRNA_intronic | 0.038 |
| rs59149647 | 6 | 116679029 | A | 0.1352 | 0.218262 | 0.0468439 | 4.45E-06 | DSE:RP1-93H18.1 | 00:00 | ncRNA_intronic | 0.124 |
| rs113865485 | 9 | 116229738 | A | 0.02087 | 0.461864 | 0.0968897 | 4.45E-06 | RGS3 | 0 | intronic | 0.127 |
| rs60749937 | 6 | 116648592 | A | 0.1332 | 0.215926 | 0.0463699 | 4.48E-06 | DSE:RP1-93H18.1 | 00:00 | ncRNA_intronic | 9.537 |
| rs57818125 | 6 | 116648060 | T | 0.1332 | 0.215863 | 0.0463715 | 4.51E-06 | DSE:RP1-93H18.1 | 00:00 | ncRNA_intronic | 9.135 |
| rs8040839 | 15 | 44118890 | T | 0.09543 | 0.257496 | 0.0550392 | 4.59E-06 | WDR76 | 270 | upstream | 3.95 |
| rs1160705 | 5 | 25128668 | T | 0.4324 | -0.165325 | 0.0363929 | 5.32E-06 | RP11-549K20.1 | 0 | ncRNA_intronic | 5.354 |
| rs116260600 | 2 | 110006345 | A | 0.0159 | -0.924414 | 0.227852 | 5.33E-06 | SH3RF3 | 0 | intronic | 1.75 |
| rs111294065 | 15 | 68575696 | C | 0.03777 | -0.559589 | 0.130372 | 5.33E-06 | FEM1B | 0 | intronic | 4.238 |
| rs147678158 | 2 | 212104028 | G | 0.01093 | 0.738721 | 0.154078 | 5.47E-06 | AC013404.1 | 59637 | intergenic | 1.812 |
| rs112889512 | 9 | 116194189 | T | 0.0328 | 0.418393 | 0.0889459 | 5.55E-06 | C9orf43 | 2224 | intergenic | 1.134 |
| rs2125950 | 8 | 2928234 | G | 0.4036 | -0.165104 | 0.0362821 | 5.84E-06 | CSMD1 | 0 | intronic | 3.327 |
| rs162253 | 17 | 3295368 | G | 0.4225 | 0.163402 | 0.0360851 | 5.87E-06 | OR1E1 | 5029 | intergenic | 2.571 |
| rs73548954 | 6 | 116644488 | T | 0.1471 | 0.211545 | 0.0460265 | 5.87E-06 | DSE:RP1-93H18.1 | 00:00 | ncRNA_intronic | 4.493 |
| rs36019747 | 1 | 186463784 | G | 0.01988 | -0.546777 | 0.128017 | 5.88E-06 | GS1-304P7.3 | 27120 | intergenic | 2.406 |
| rs11113754 | 12 | 108525445 | G | 0.2217 | 0.190136 | 0.0415799 | 6.08E-06 | WSCD2 | 0 | intronic | 3.834 |
| rs11113755 | 12 | 108525533 | T | 0.2217 | 0.19014 | 0.0415821 | 6.08E-06 | WSCD2 | 0 | UTR5 | 12.56 |
| rs11113756 | 12 | 108525595 | G | 0.2217 | 0.190143 | 0.0415831 | 6.08E-06 | WSCD2 | 0 | UTR5 | 6.547 |

**Caption**

Annotation of variants was performed by FUMA tool (<https://fuma.ctglab.nl/>).

**Abbreviations**

chr = chromosome; dist = distance to the nearest gene; EA = effect allele; func = functions annotated from ANNOVAR; MAF = minor allele frequency; MDD = major depressive disorder; pos = position on hg19; rsid = variant ID; SE = standard error; SSRI = selective serotonin reuptake inhibitors.

### Supplementary Figures

#### Supplementary figure 1. Schematic diagram to illustrate (A) a switcher from paroxetine (SSRI) to clomipramine (TCA); (B) a non-switcher for fluoxetine (SSRI).

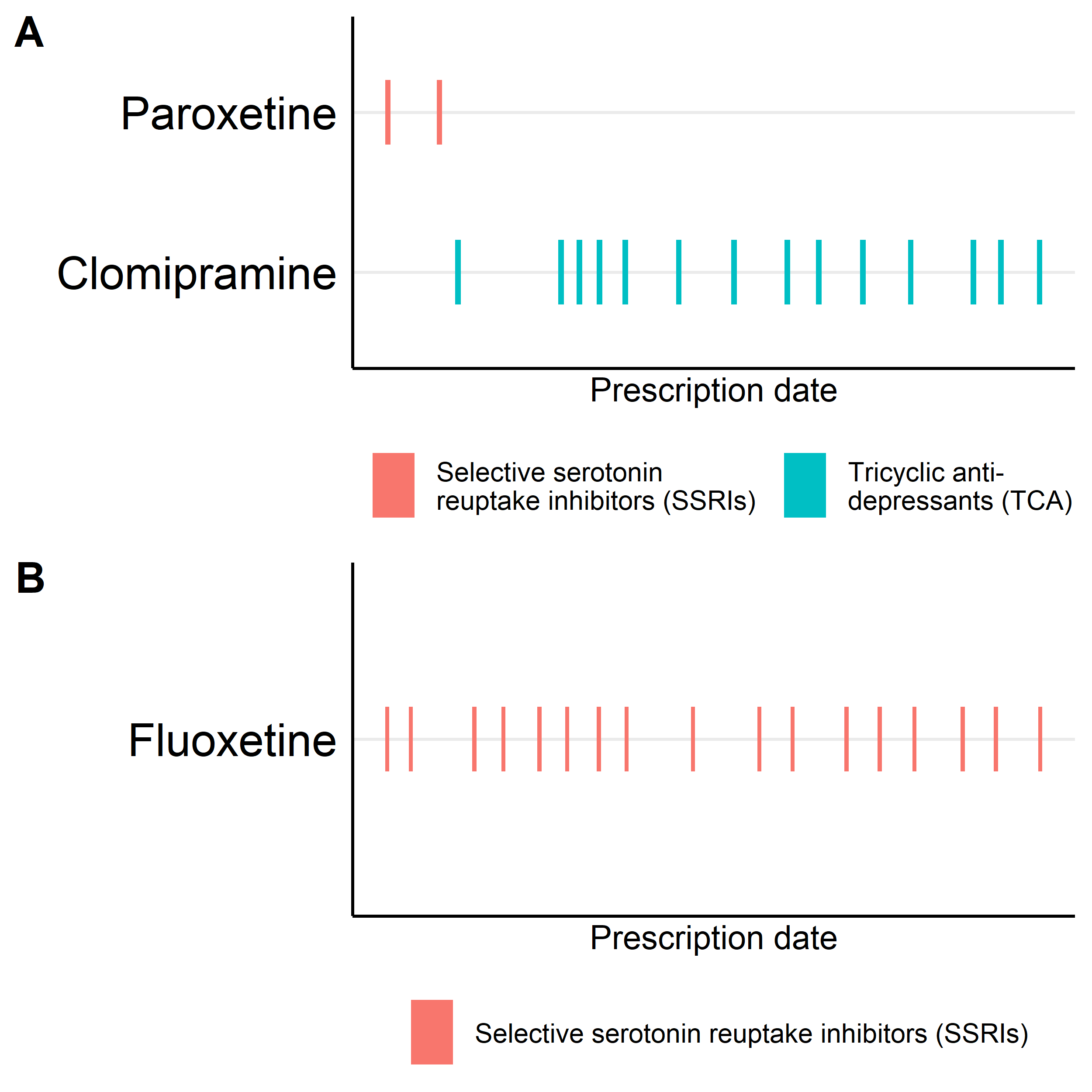

**Abbreviations**

SSRI = selective serotonin reuptake inhibitors; TCA = tricyclic antidepressants.

#### Supplementary figure 2. Sample sizes for SSRI switching, stratified by diagnosis criteria

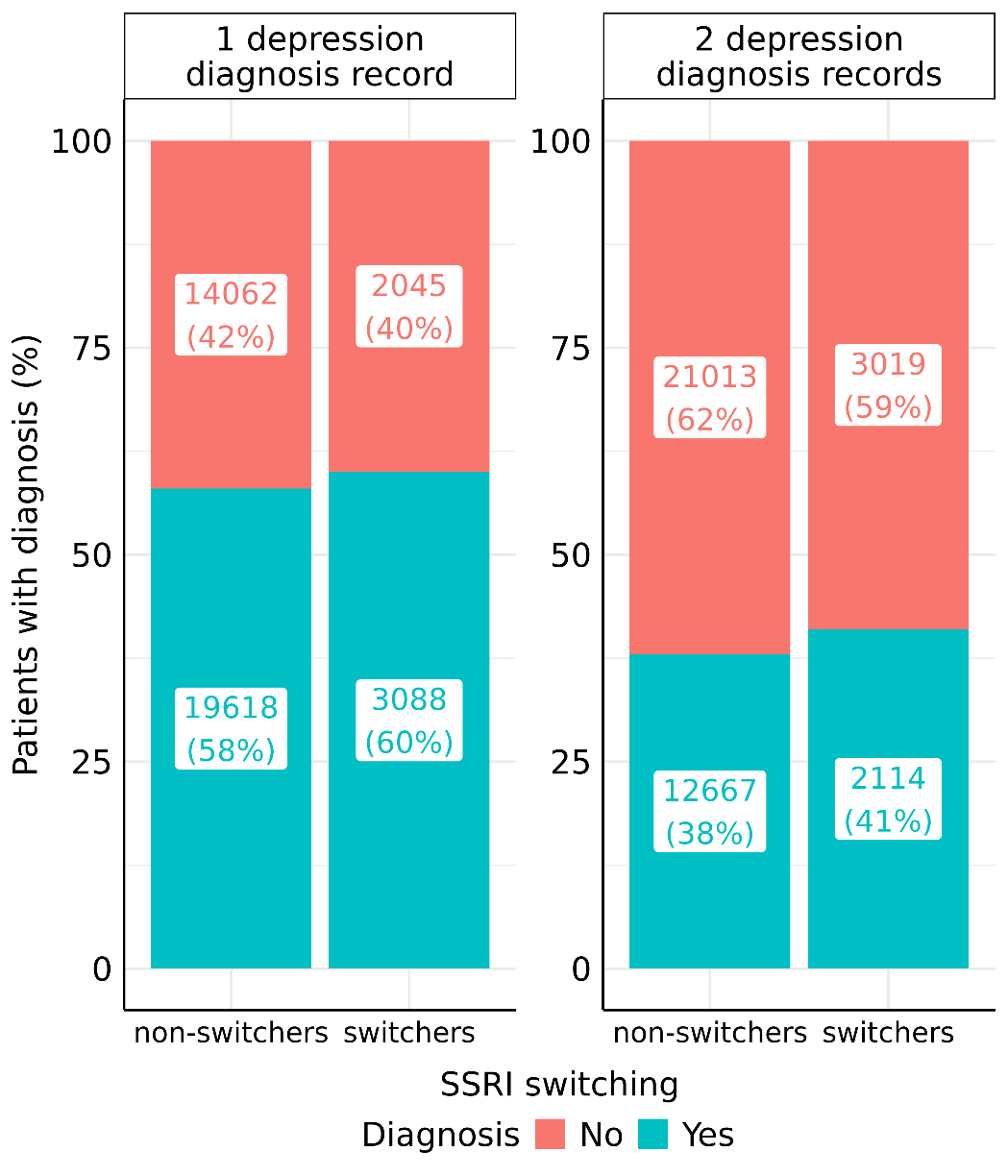

**Caption**

Numbers expressed as count (percentage).

**Abbreviations**

SSRI = selective serotonin reuptake inhibitors.

#### Supplementary figure 3. Distribution of prescriptions in primary sample, UK Biobank

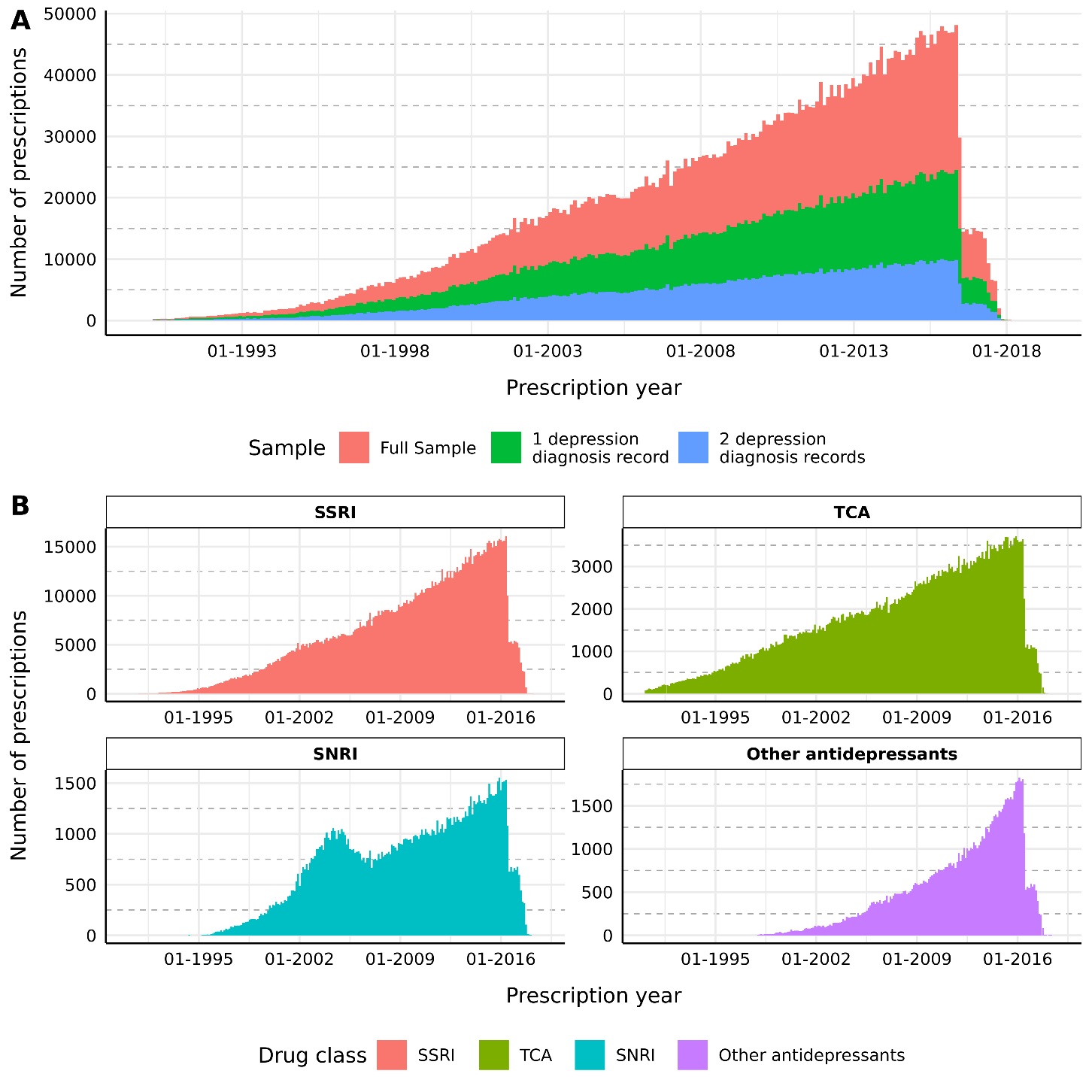

**Caption**

**(A)** Number of prescriptions across time stratified by sample; **(B)** Number of prescriptions in full primary sample stratified by 4 antidepressant classes with most prescriptions.

Histogram constructed by dividing prescriptions into 250 bins according to date of prescription.

The decrease in SSRI prescriptions in 2016 is due to different data extraction dates for different providers of EHR systems to UKB, ranging from August 2016 to July 2017.

**Abbreviations**

SNRI = serotonin–norepinephrine reuptake inhibitors; SSRI = selective serotonin reuptake inhibitors; TCA = tricyclic antidepressants.

#### Supplementary figure 4. Distribution plot for time to switch for SSRI switchers in primary sample, UK Biobank

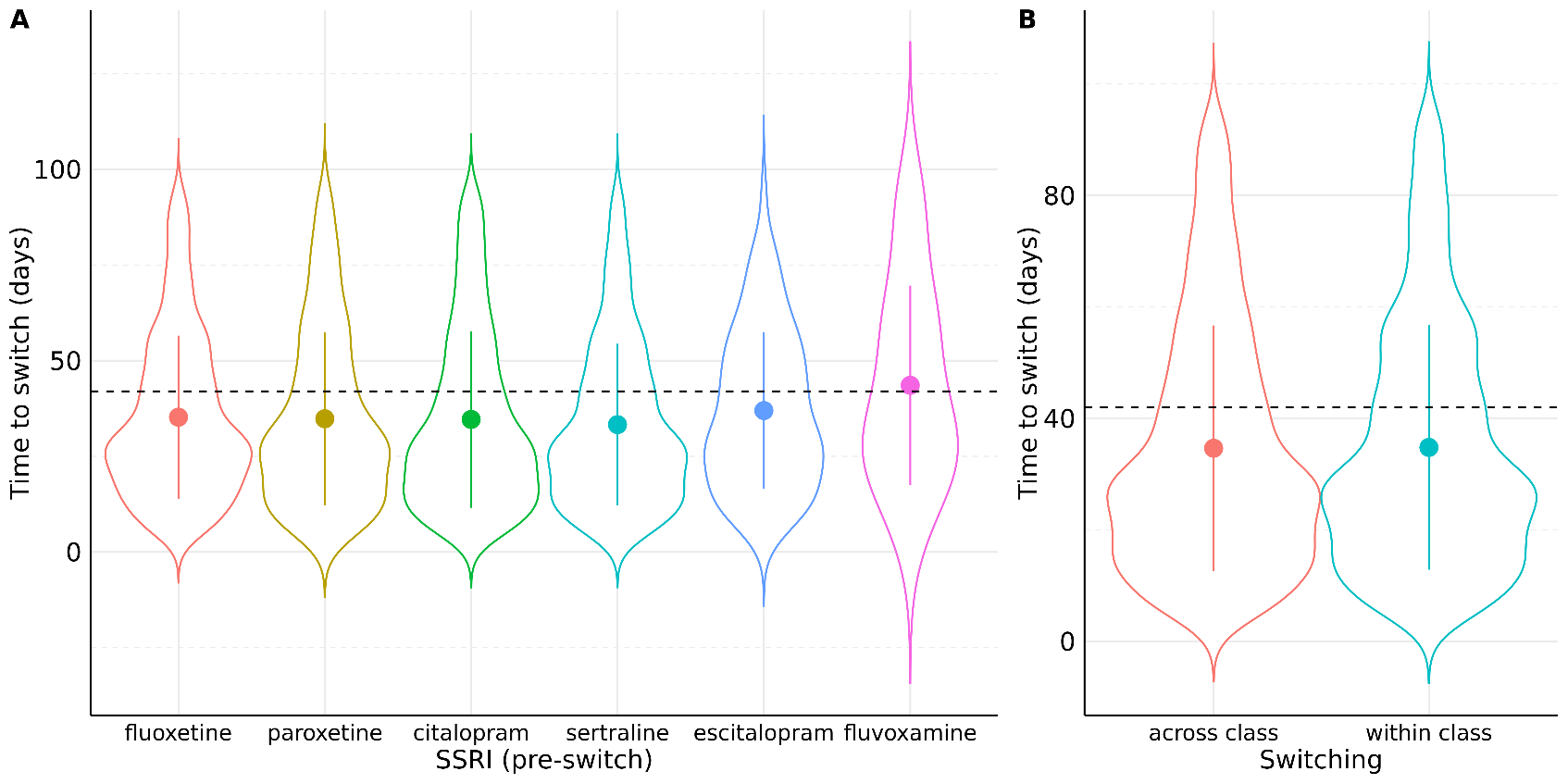

**Caption**

Distribution plots for **(A)** index SSRI before switching; **(B)** comparison between across-class (other drug classes) and within-class switches (to another SSRI). Dashed lines (black) represent 42 days (6 weeks) from index prescription of SSRI.

**Abbreviations**

SSRI = selective serotonin reuptake inhibitors.

#### Supplementary figure 5. Distribution of index SSRI used and age at index date for SSRI prescription of primary sample, UK Biobank

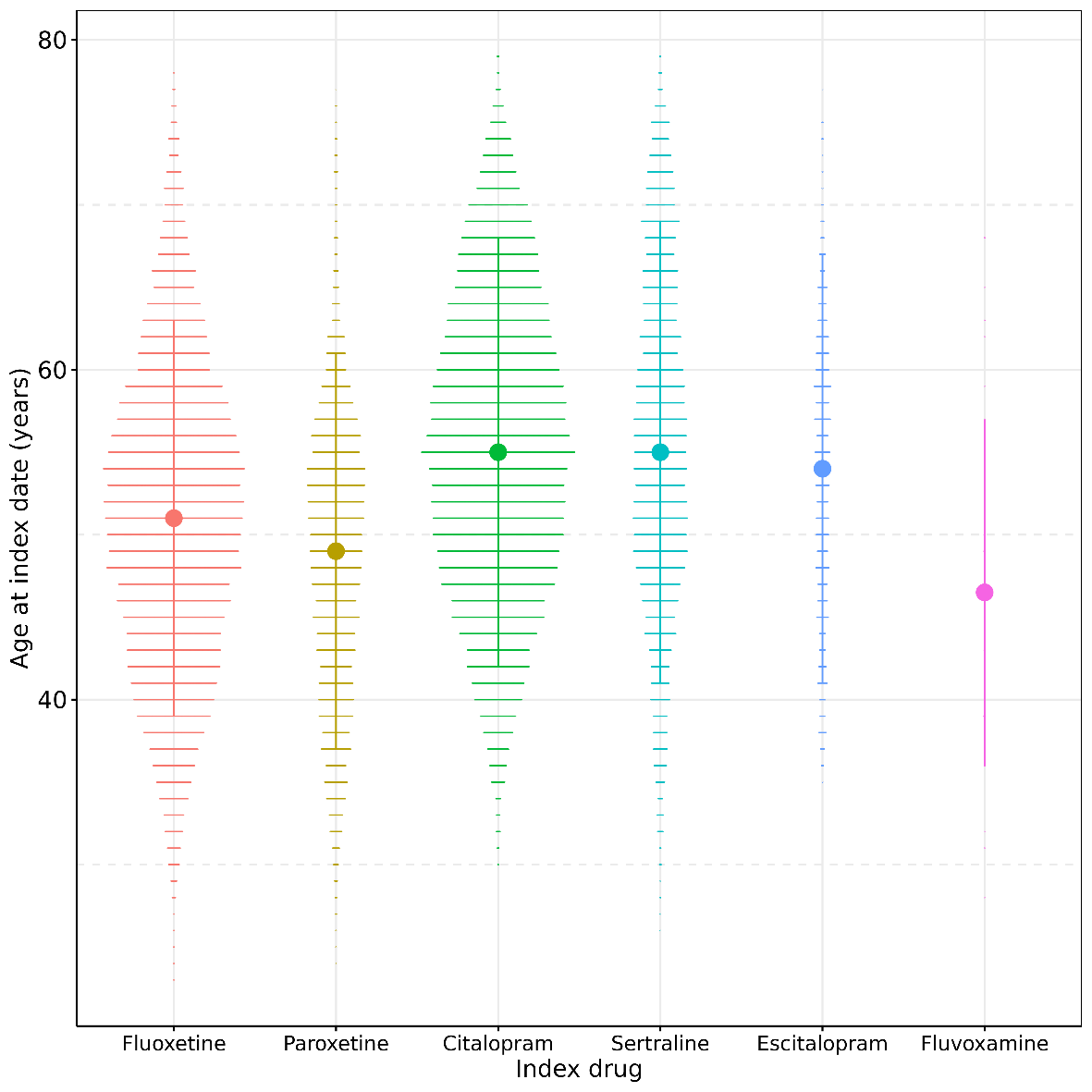

**Caption**

Index drug defined as: (1) switchers: SSRI that is switched subsequently; (2) non-switchers: SSRI with ≥ 3 consecutive prescriptions.

**Abbreviations**

SSRI = selective serotonin reuptake inhibitors.

#### Supplementary figure 6. Sex-specific descriptive analysis for primary sample, UK Biobank

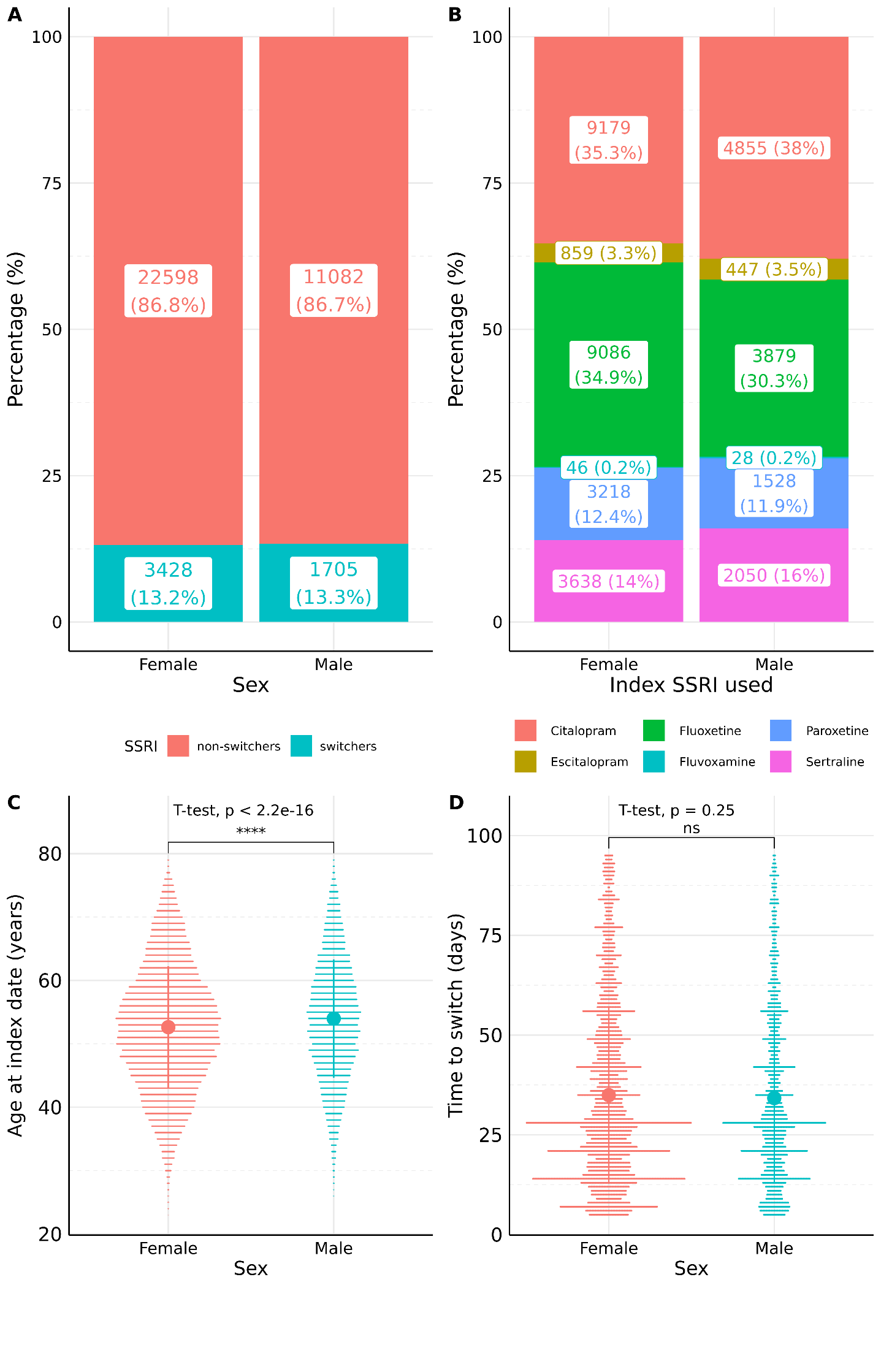

**Caption**

Sex-specific analysis for **(A)** proportion of switchers; **(B**) index SSRI used; **(C)** age at index date (years); **(D)** time to switch (days).

Proportion of switchers and index SSRI used expressed in count (percentage).

Statistical significance for gender-specific differences in mean age at index date and time to switch assessed by T-test (two-sided).

**Abbreviations**

ns = not-significant; SSRI = selective serotonin reuptake inhibitors.

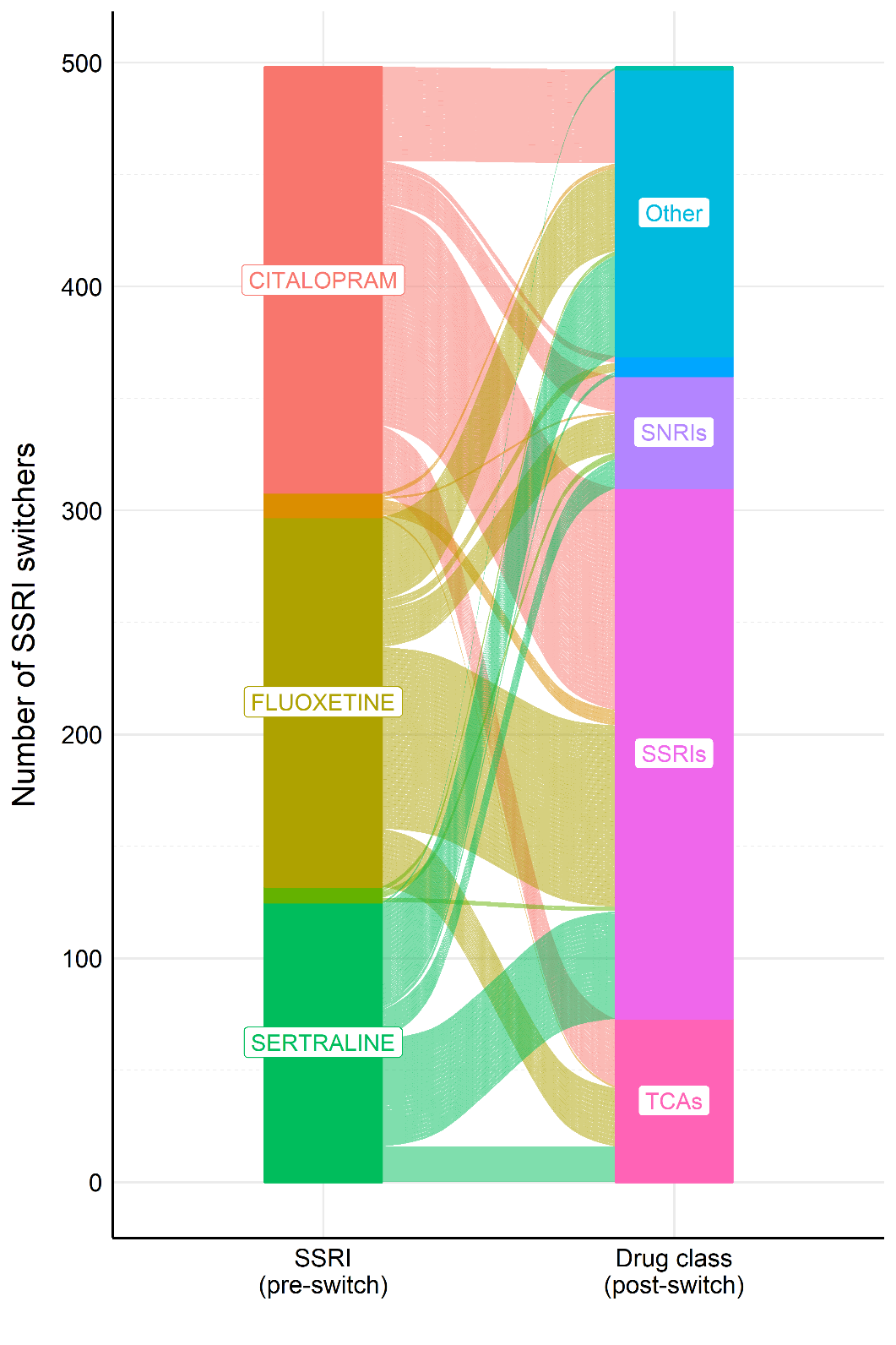

#### Supplementary figure 7. Alluvial plot for switching patterns in Generation Scotland replication sample

**Abbreviations**

SNRIs = serotonin–norepinephrine reuptake inhibitors; SSRIs = selective serotonin reuptake inhibitors; TCAs = tricyclic antidepressants.

#### Supplementary figure 8: Distribution of switch patterns across assessment centres

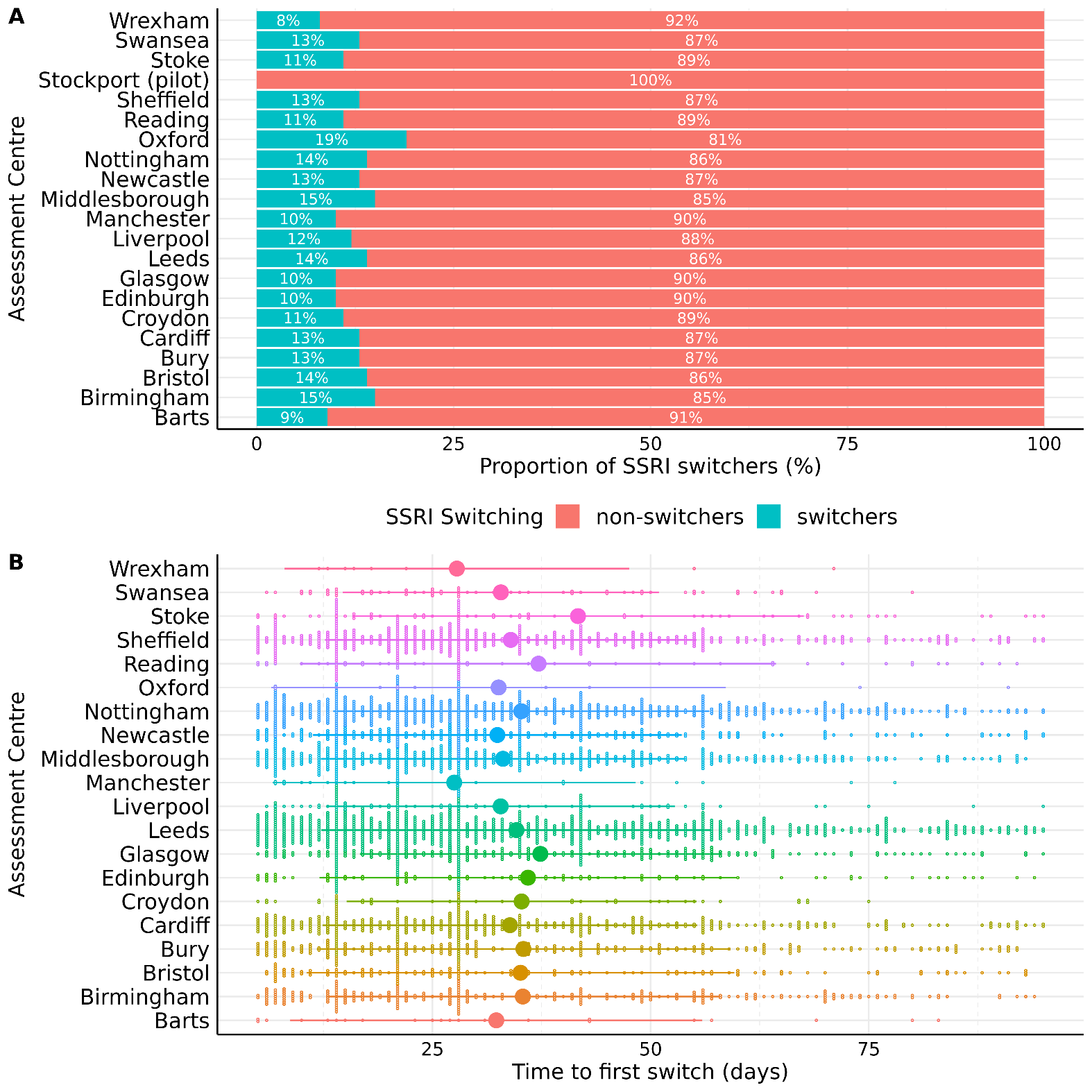

**Abbreviations**

SSRI = selective serotonin reuptake inhibitors.

#### Supplementary figure 9. Relationship between switching patterns and SSRI index date percentiles in UKB

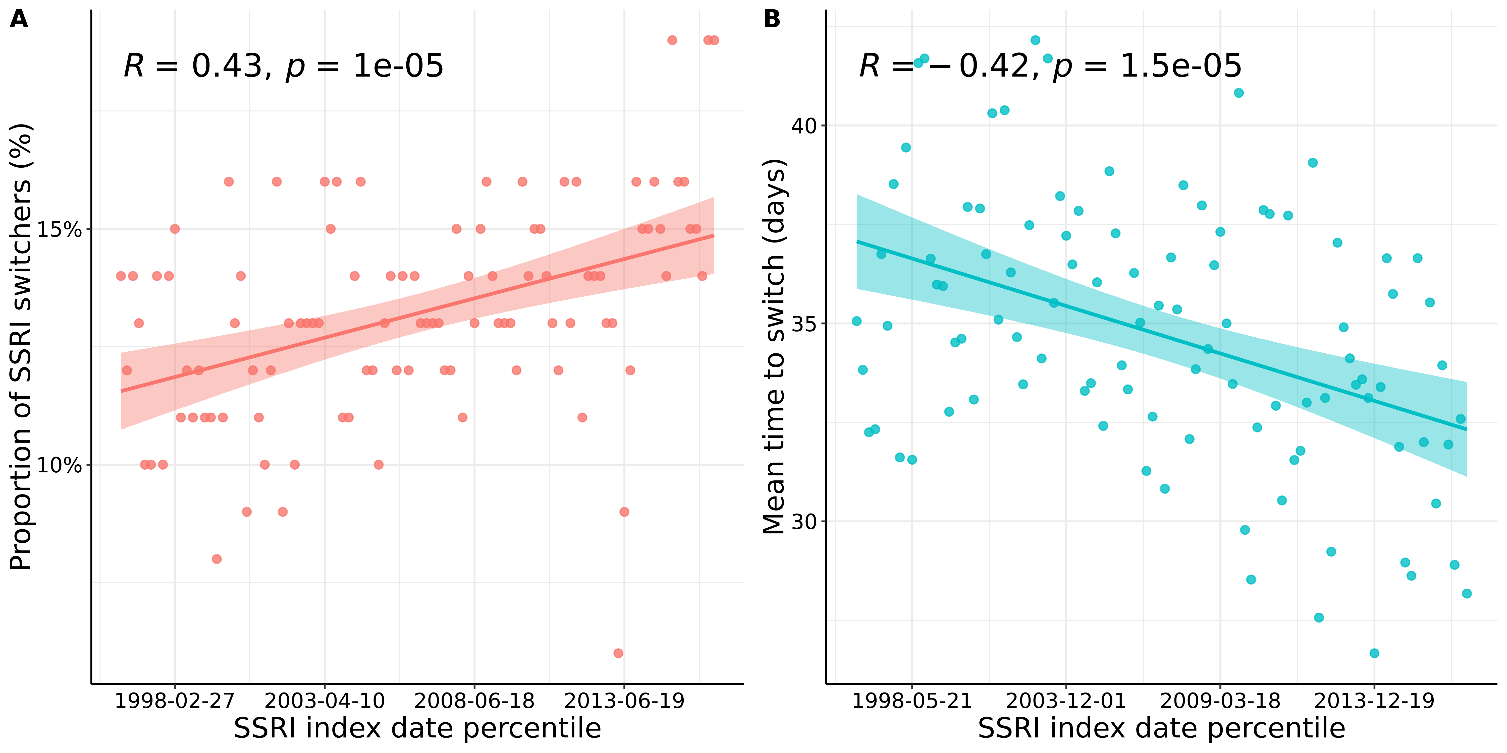

**Caption**

Correlation plots between **(A)** proportion of SSRI switchers within each percentile; **(B)** mean time to switch (days) within each percentile, and index date percentile of SSRI.

**Abbreviations**

SSRI = selective serotonin reuptake inhibitors.

#### Supplementary figure 10. BMI distribution at assessment for primary sample, UK Biobank

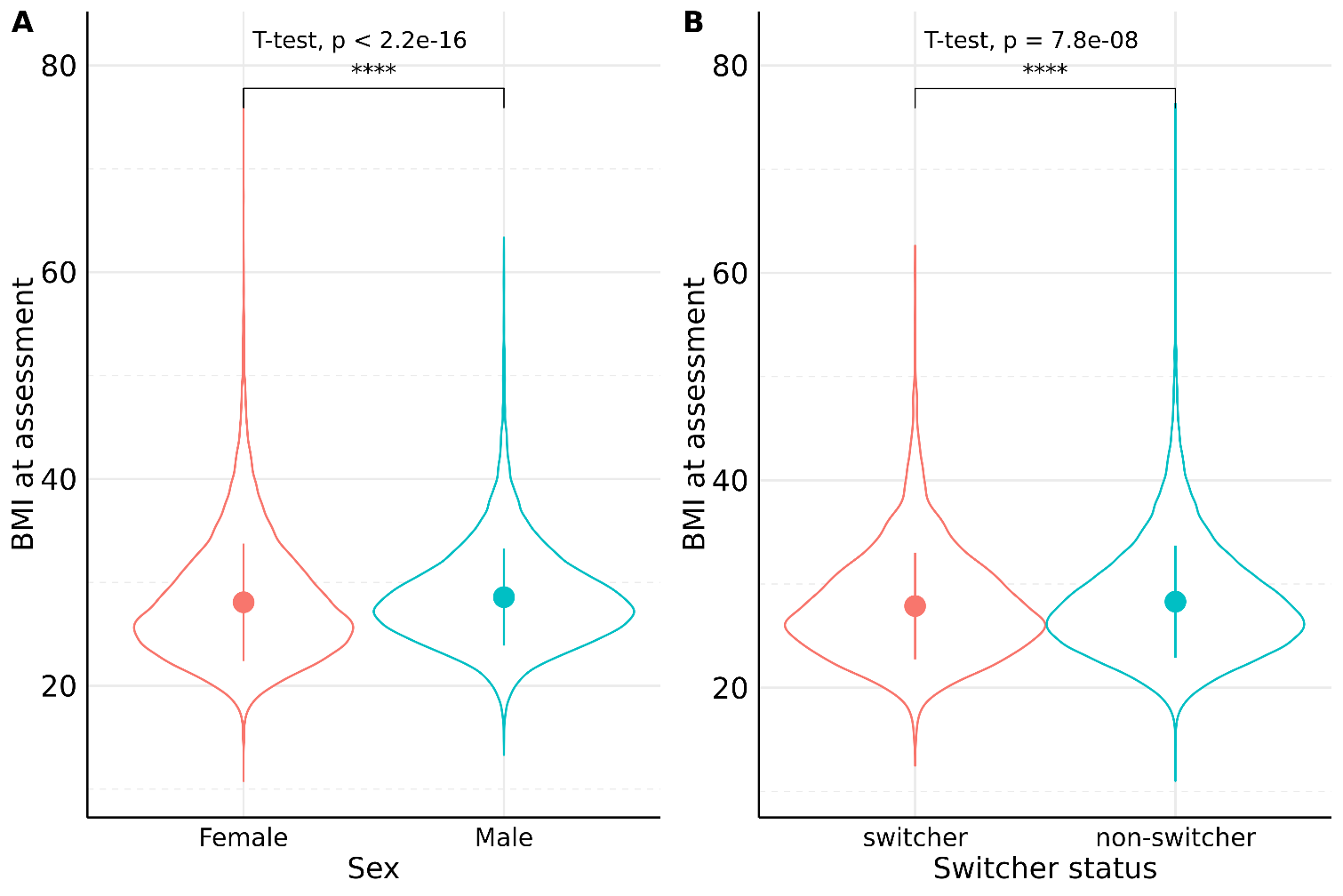

**Caption**

Statistical significance in mean between groups assessed by two-sided t-test.

Dots and error bars represent mean and standard deviations of distributions respectively.

**Abbreviations**

BMI = body mass index.

#### Supplementary figure 11: Distribution of neuroticism scores at assessment for primary sample

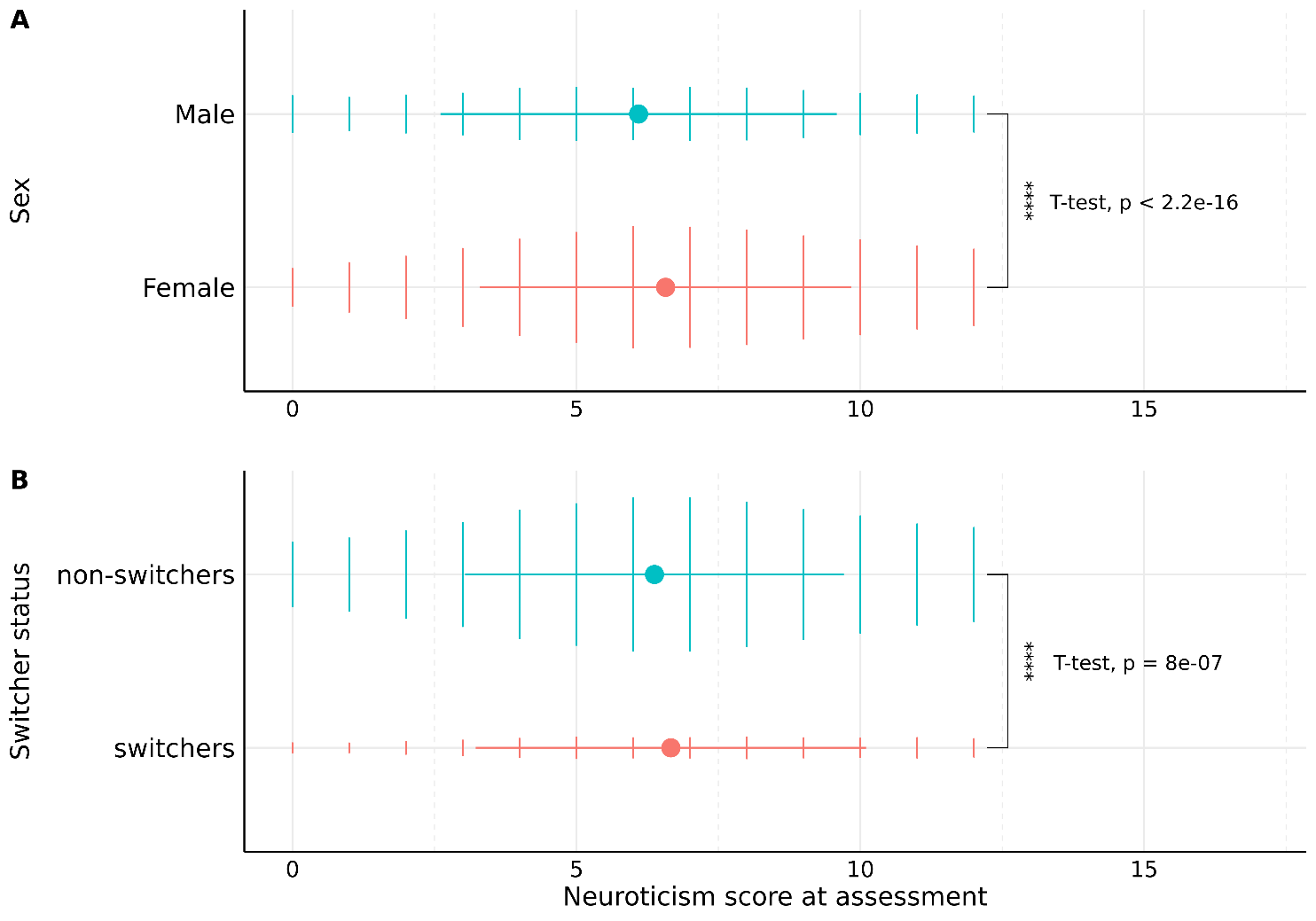

**Caption**

Statistical significance in mean between groups assessed by two-sided t-test.

Dots and error bars represent mean and standard deviations of distributions respectively.

#### Supplementary figure 12. Distribution of demographic variables across assessment centres

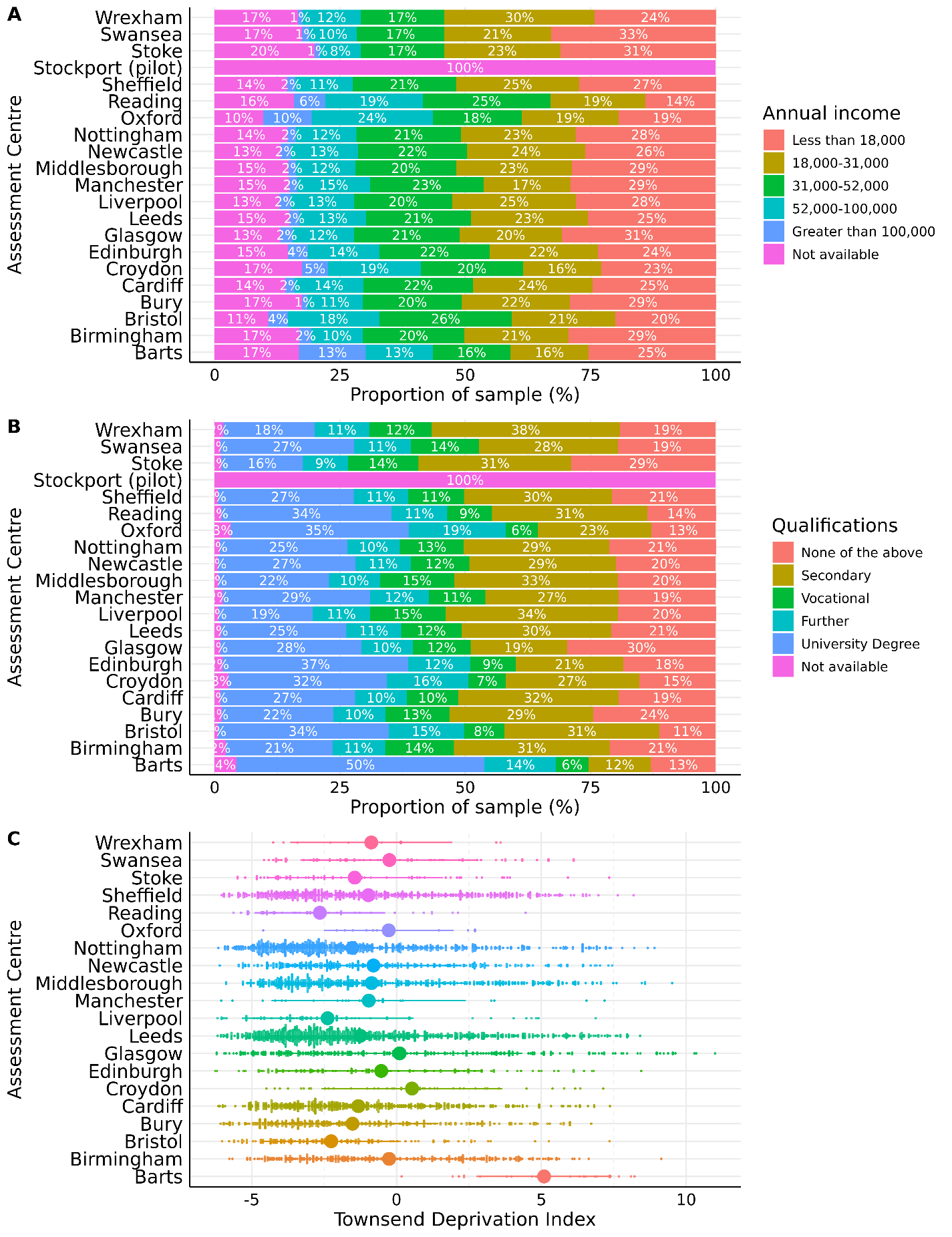

#### Supplementary figure 13: GCTB estimates on *Pi* (polygenicity) and *S* (negative selection)

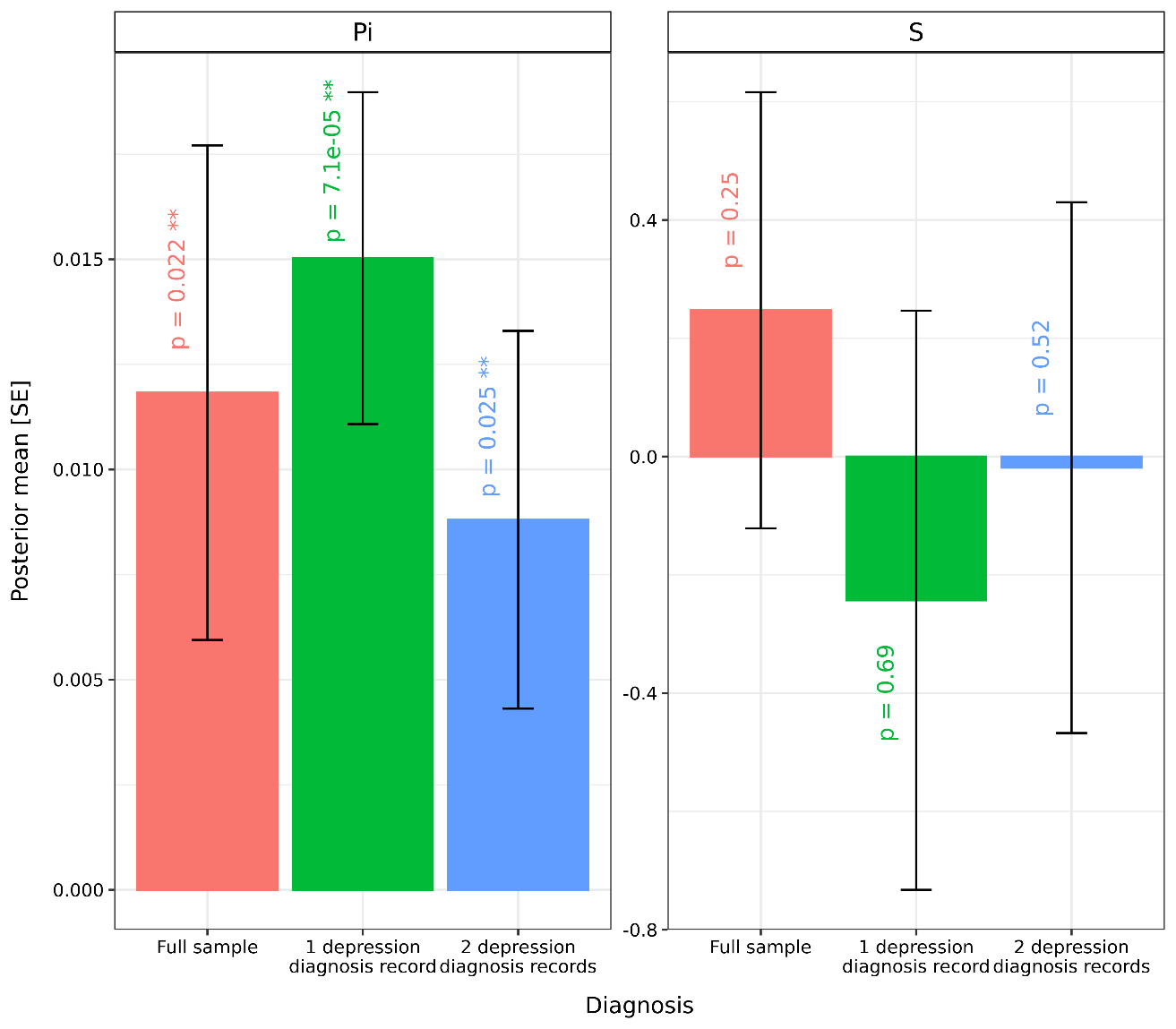

**Caption**

GCTB estimates on number of Pi (degree of polygenicity) and S (value for natural selection), expressed as posterior mean (standard error).

**Abbreviations**

GCTB = Genome-wide Complex Trait Bayesian; SE = standard error.

#### Supplementary figure 14. Heritability distribution for GCTB per iteration in MCMC sampler

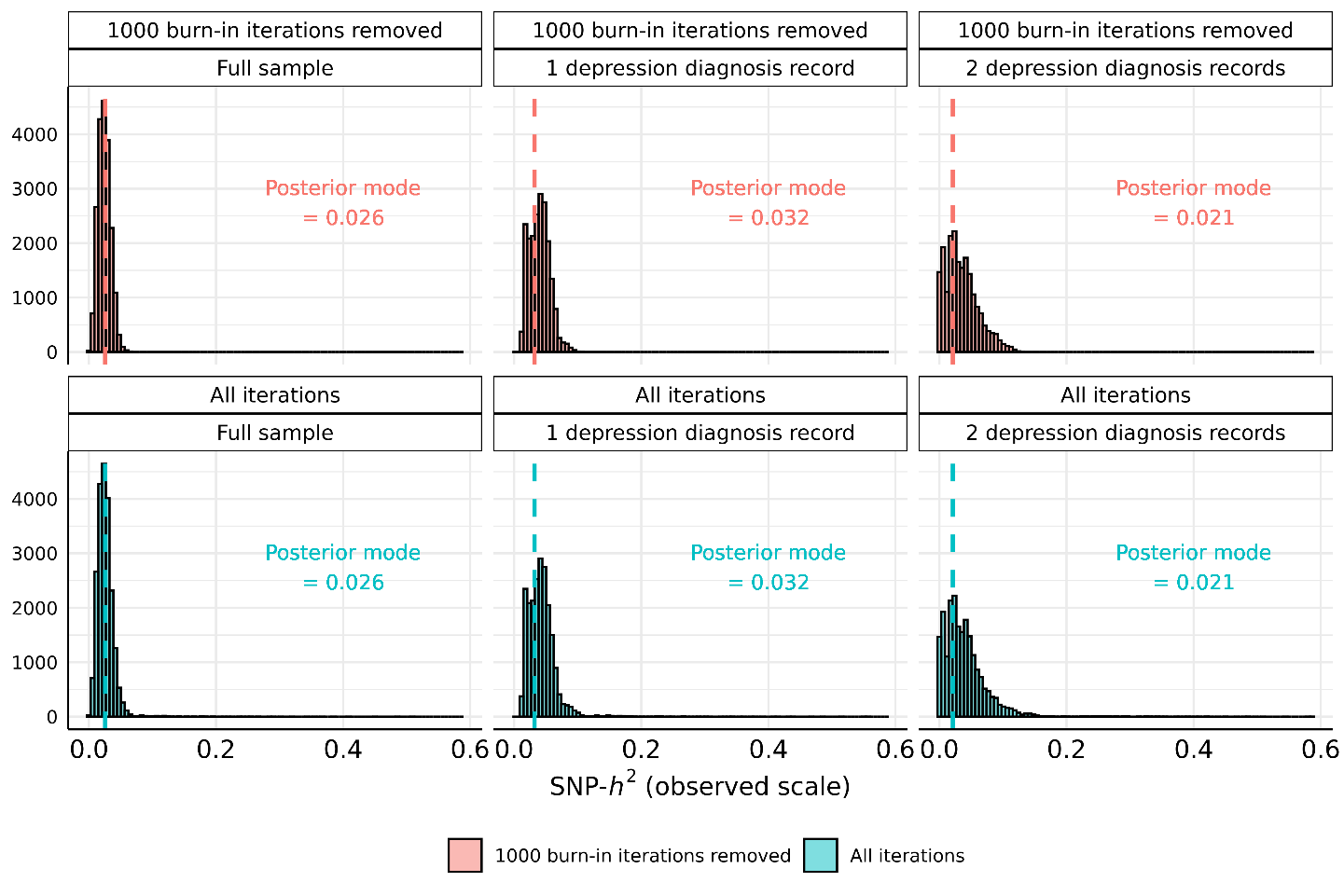

**Caption**

Heritability estimates divided into 100 bins for visualization. 21000 iterations (with 1000 burn-in iterations removed) were used as the options for MCMC sampling in GCTB estimation of heritability.

**Abbreviations**

GCTB = Genome-wide Complex Trait Bayesian; *h^2^* = heritability; MCMC = Markov chain Monte Carlo; SNP = single-nucleotide polymorphism.

#### Supplementary figure 15. Manhattan plot for SSRI switching (full sample)

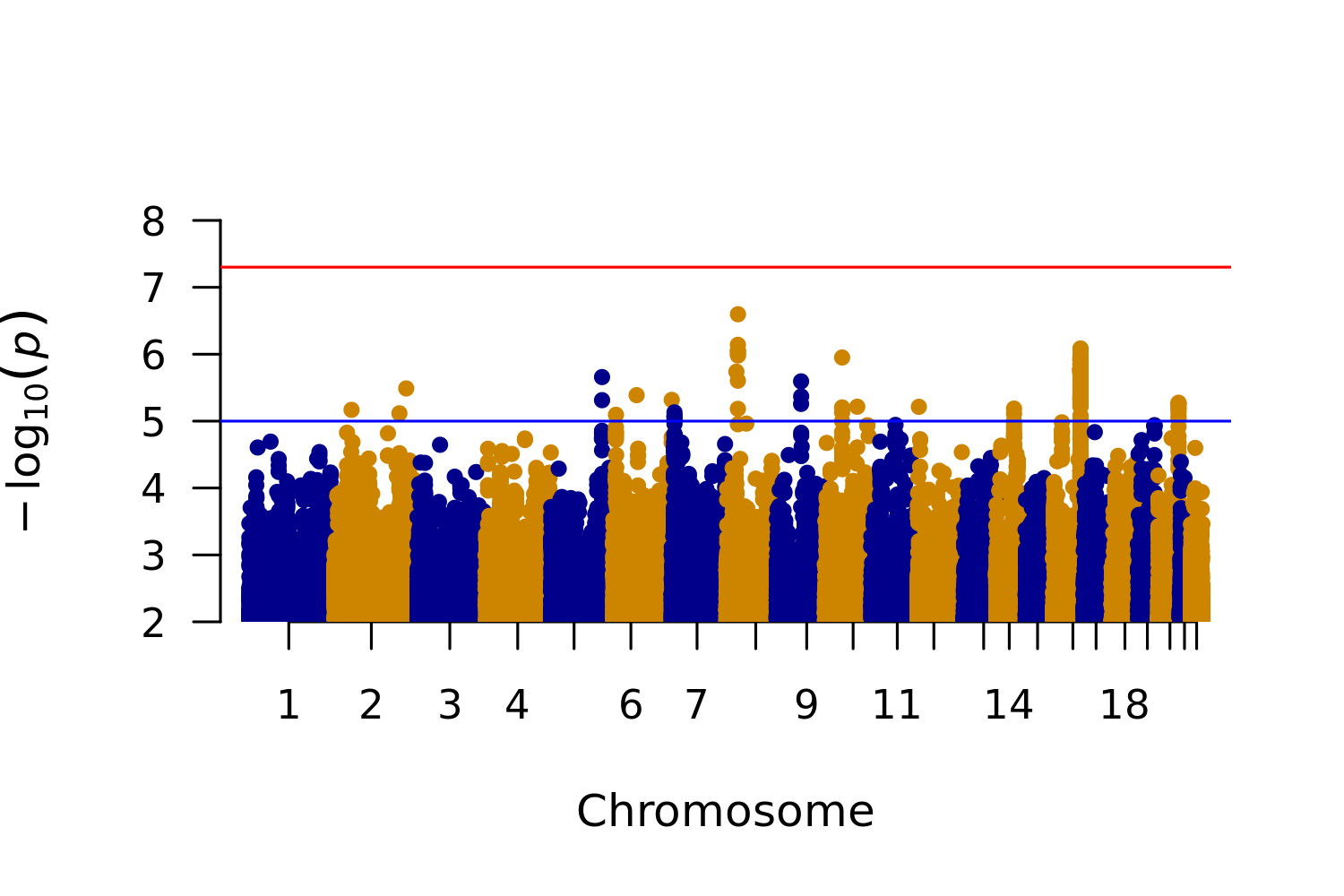

**Caption**

Red line represents genome-wide significance (p < 5 × 10^-8^). Blue line represents suggestive hits (p < 1 × 10^-5^).

**Abbreviations**

SSRI = selective serotonin reuptake inhibitors.

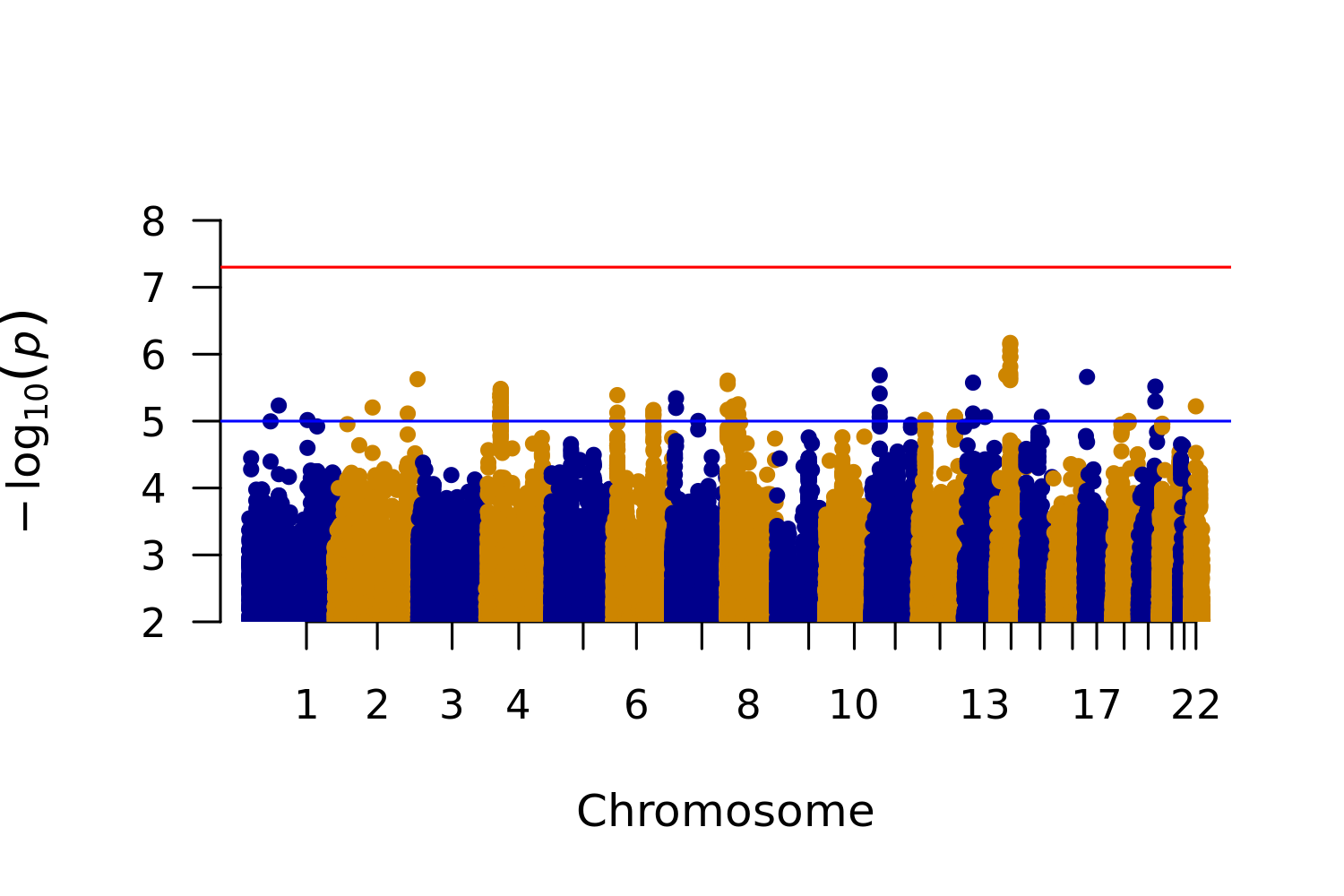

#### Supplementary figure 16: Manhattan plot for SSRI switching (≥ 1 depression diagnosis record)

**Caption**

Red line represents genome-wide significance (p < 5 × 10^-8^). Blue line represents suggestive hits (p < 1 × 10^-5^).

**Abbreviations**

MDD = major depressive disorder; SSRI = selective serotonin reuptake inhibitors.

#### Supplementary figure 17: Manhattan plot for SSRI switching (≥ 2 depression diagnosis records)

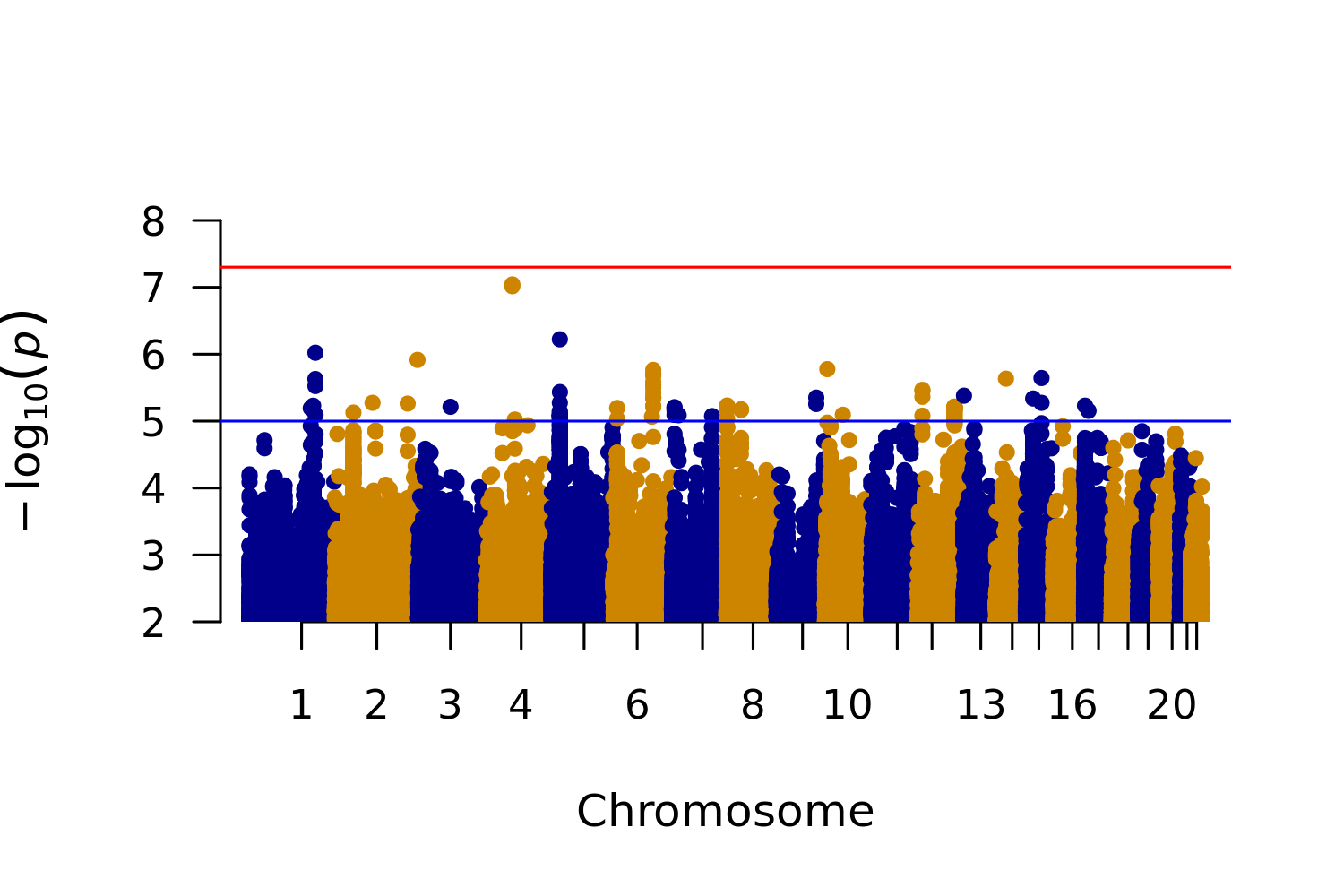

**Caption**

Red line represents genome-wide significance (p < 5 × 10^-8^). Blue line represents suggestive hits (p < 1 × 10^-5^).

**Abbreviations**

MDD = major depressive disorder; SSRI = selective serotonin reuptake inhibitors.

#### Supplementary figure 18. QQ plot for SSRI switching (full sample)

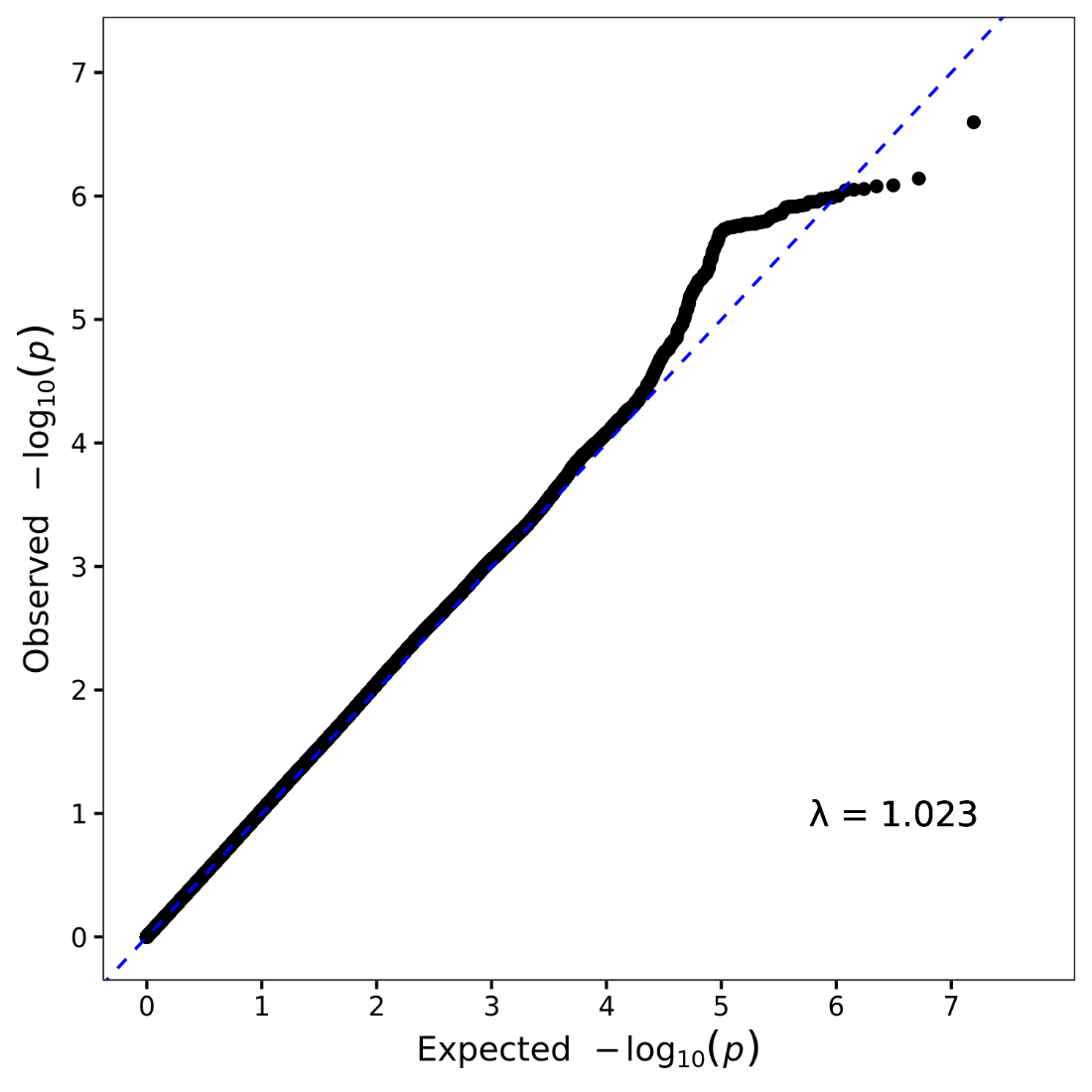

**Abbreviations**

λ = lambda for genomic control; MDD = major depressive disorder; SSRI = selective serotonin reuptake inhibitors.

#### Supplementary figure 19. QQ plot for SSRI switching (≥ 1 depression diagnosis record)

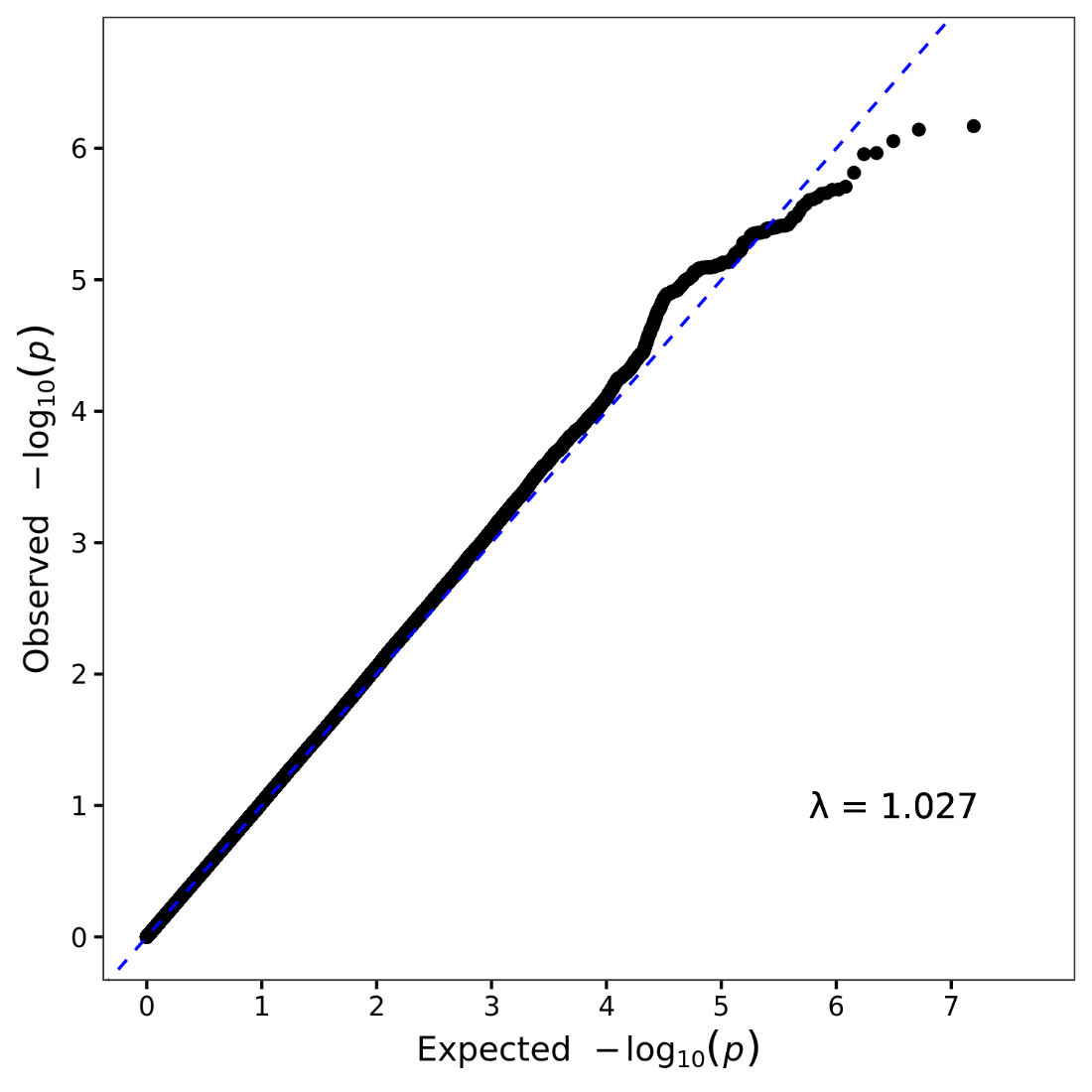

**Abbreviations**

λ = lambda for genomic control; MDD = major depressive disorder; SSRI = selective serotonin reuptake inhibitors.

#### Supplementary figure 20. QQ plot for SSRI switching (≥ 2 depression diagnosis records)

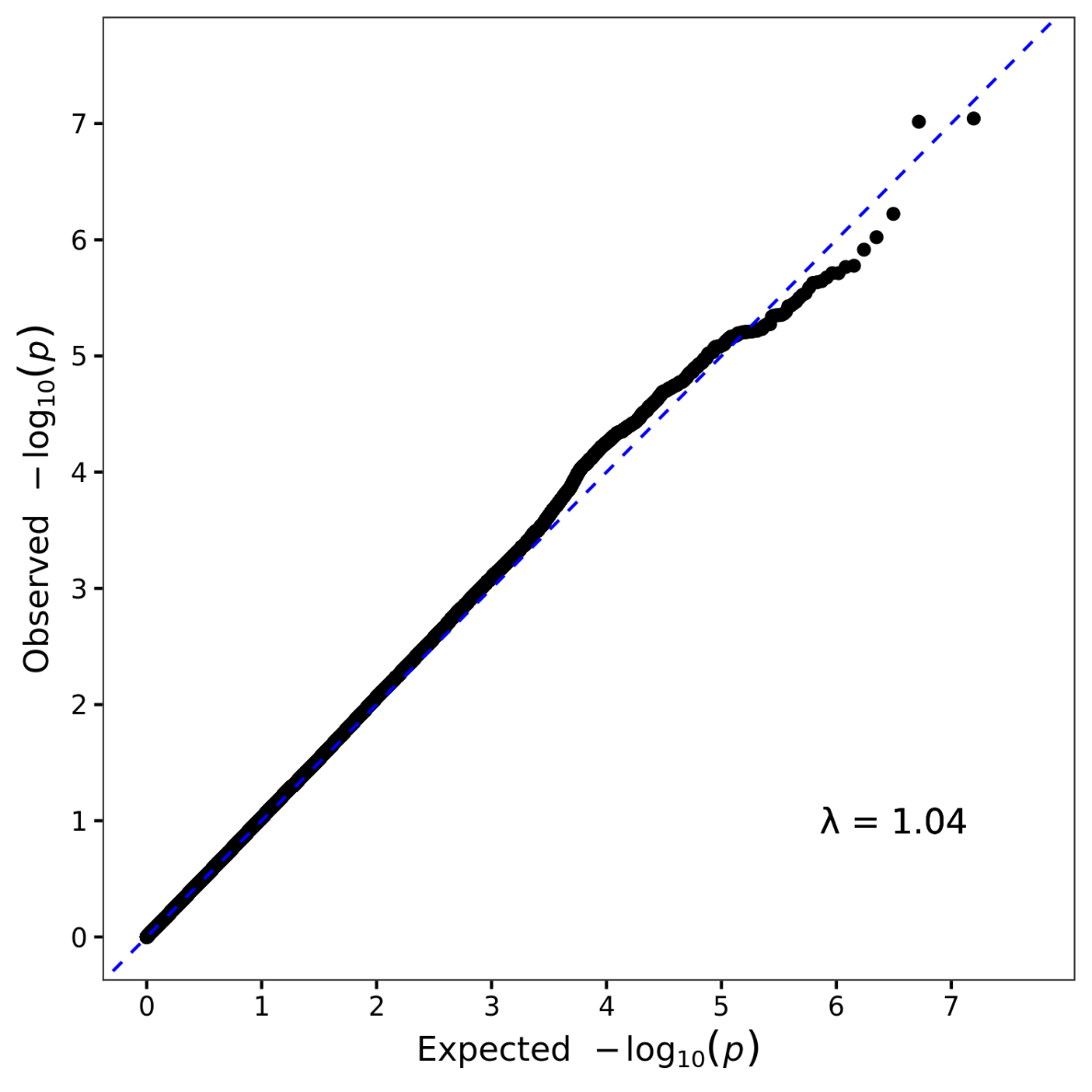

**Abbreviations**

λ = lambda for genomic control; MDD = major depressive disorder; SSRI = selective serotonin reuptake inhibitors.
